# Prediction of Cardiovascular and Renal Complications of Diabetes by a multi-Polygenic Risk Score in Different Ethnic Groups

**DOI:** 10.1101/2025.06.17.25329804

**Authors:** Edoh Kodji, Redha Attaoua, Mounsif Haloui, Camil Hishmih, Mirjam Seitz, Pavel Hamet, Julie Hussin, Johanne Tremblay

## Abstract

We developed a multi-Polygenic risk score (multiPRS) to predict the risk of nephropathy, stroke, and myocardial infarction in people with type 2 diabetes of European descent. The underrepresentation of non-European populations remains a major challenge in genomics research. **Objective**: To evaluate the ability of our multiPRS model to accurately predict these complications in patients of African and South Asian descents. **Method**: The multiPRS was developed using 4098 participants with type 2 diabetes of European origin from the ADVANCE trial. Its predictive performance was tested on 17,574 White British, 1,145 South Asian and 749 African participants with type 2 diabetes from the UK Biobank using different machine learning prediction models, including techniques tailored for imbalanced datasets. **Results**: Globally, linear discriminant analysis and logistic regression had the best performance to predict the risk of nephropathy, stroke, and myocardial infarction in people with type 2 diabetes for the three ethnic groups. Mondrian Cross-Conformal Prediction method when added to logistic regression improved the AUROC values and case detection, particularly in South Asians and Africans, while in White British, performance varied by phenotype. **Conclusion**: Logistic regression, when used as the underlying model within the Modrian Cross-Conformal Prediction framework, improved the prediction performance, with a confidence level, of diabetes complications and allows better translation of a multiPRS derived from European populations to other ethnic groups.

## Introduction

Diabetes burden increases worldwide, and all ages, continents and communities are affected. In 2021, worldwide, 537 million adults lived with diabetes (prevalence: 10 %). The number of people with diabetes is expected to rise to 783 million by 2045[1]. Type 2 diabetes (T2D) accounts for 90-95% of all diabetes cases[2]. T2D is associated with serious complications, including cardiovascular disease (CVD) and renal failure, which significantly contribute to patient morbidity and healthcare burden. Early identification of risk prior to development of complications is crucial to enable targeting individuals that could benefit from early or novel therapeutic interventions[3, 4].

In recent years, polygenic risk scores (PRS) have emerged as promising tools to predict susceptibility to complex diseases like T2D and its complications [5]. However, most PRS have been developed from individuals of European ancestry, raising concerns about their generalizability across diverse populations [6–11]. Key challenges include differences in genetic architecture, sample size limitations, population admixture, and the underrepresentation of non-European groups in genetic databases. While creating PRS tailored to diverse ethnic groups is theoretically ideal, it is currently impractical due to data scarcity and the high costs associated with large-scale genome-wide association studies (GWAS) in underrepresented populations. Therefore, alternative strategies are needed to improve the transferability of existing PRS models, particularly those developed in European cohorts, to other ancestral groups.

We previously developed a multi-polygenic risk score (multiPRS) combining ten weighted PRS (wPRS) for outcomes and risk factors associated to T2D. The multiPRS stratified T2D patients according to risk of cardiovascular and renal complications. Our study also demonstrated that participants identified at high-risk by our multiPRS were those who benefited most from the intensive therapy administered in ADVANCE trial [5, 12]. The predictive model was initially developed in individuals with T2D of European (EUR) descent and its performance in other populations is unknown. Here, we assessed whether this model could be adapted to accurately predict diabetes complications in two additional major ethnic groups. We compared different machine learning prediction models, including techniques tailored for imbalanced datasets.

Mondrian Cross-Conformal Prediction (MCCP) has gained attention in biomedical and genomic modeling for its ability to quantify prediction confidence under class imbalance. This approach evaluates the “nonconformity” of a new instance by comparing it to a training set, using a nonconformity measure (NCM). Conformal prediction provides a prediction region that can include multiple possible labels with some specified confidence level, ensuring overall statistical validity [13–16]. MCCP combines Mondrian inductive conformal prediction with a cross-calibration procedure to estimate prediction confidence on imbalanced datasets [15, 17]. Therefore, to improve the reliability of individual-level predictions in the context of population transferability and imbalanced data, we explored the use of MCCP, enabling us to identify not only likely cases and controls, but also individuals for whom the model is uncertain, a critical feature in clinically actionable risk stratification.

## Ethics

The project was approved by the ethics committee of the Centre hospitalier de l’Université de Montréal (CHUM) under the number 2023-11126,22.205-LM.

## Methods

Our multiPRS predictive model was based on 598 SNPs identified by summary statistics of meta-analyses of publicly available genome-wide association studies (GWAS) gathering genomic and phenotypic information from over one million individuals of European descent[12]. We combined 10 weighted polygenic risk scores (PRS) gathering genomic variants associated to cardiovascular and renal diseases alongside their key risk factors, the first four principal component of ethnicity, sex, age at onset and diabetes duration into one logistic regression (LR) model, to predict renal and cardiovascular complications of T2D in a single prediction model using samples and data from European participants of T2D of the ADVANCE trial[12, 18]. Genotyping and imputation of the ADVANCE samples were performed as described in[5]. Details on the construction and the multi-PRS model and list of SNPs are available in the Supplementary Material of our previous study [5].

## Cohorts

**ADVANCE** (NCT 00145925) was a 2×2 factorial design, randomized controlled trial of blood pressure (BP) lowering (perindopril-indapamide *vs* placebo) and intensive glucose control (gliclazide MR-based intensive intervention with a target of 6.5 HbA1c *vs* standard care) in patients with T2D. A total of 11,140 participants were recruited from 215 centers in 20 countries from Asia, Australia, North America and across Europe. They were older than 55 years and diagnosed with T2D after the age of 30 years. We used a subset of 4,098 genotyped T2D patients of genetically determined European descent followed for a period of 4.5 years in ADVANCE[12].

**UK Biobank**: This study was performed under projects 49731 and 59642. The UK Biobank study started in 2006 and, until 2010, recruited more than 500,000 participants from the general UK population, aged between 40-69[19]. Individuals had type 2 diabetes diagnosed by a doctor (Field ID 2443) at the age of 30 years or older to avoid potential type1 diabetes. We included 17,574 individuals with T2D of self-declared White British (WB) origin and 1,145 of South-Asian (SA) and 749 of African (AFR) origins living in UK whose ethnicities were confirmed by genetic ethnic grouping (Field ID 22006). Phenotypic data collected included year of birth (Field ID 34), age at recruitment (Field ID 21022), sex (Field ID 31), genetic sex (Field ID 22001), age diabetes diagnosed (Field ID 2976), age when attended assessment centre (Field ID 21003). These last 2 phenotypes allowed us to calculate the duration of diabetes (derived phenotype). eGFR, calculated by the CKD-Epi formula, used plasma creatinine levels (Field ID 30510). Albuminuria (Field ID 30500) and creatininuria (Field ID 30700) were used to calculate micro and macroalbuminuria. For systolic blood pressure, we used the automated reading (Field ID 4080), same for diastolic blood pressure (Field ID 4079), medications blood pressure or diabetes or exogenous hormones (Field ID 6153), and medications for cholesterol, blood pressure or diabetes (Field ID 6177) were used to define hypertensive participants. The samples from the UKBB were genotyped and imputed as described in our publication[5]. Around 35,000 SNPs have been used to calculate principal components (PCs) for each patient as previously reported [5]. PCs vectors are then exploited as covariates to build PRS.

## Statistical and machine learning-based prediction models

Various statistical methods, both traditional logistic regression (LR) models and modern machine learning algorithms, were used to evaluate the predictive performance of the multiPRS for each outcome. For this purpose, we used the PyCaret program, an open-source machine learning library in Python[20]. The cohort used to train the models was ADVANCE, while each of the UK Biobank populations (White British, South Asian, and African) was used separately for testing. A 5-fold cross-validation procedure was applied during model training and calibration. The input features included in the analysis were the 10 weighted PRS (wPRS), the age of T2D diagnosis, diabetes duration, patient sex, as well as the first four principal components (PC 1-4) of genetic ancestry. The statistical and machine learning methods tested are: CatBoost Classifier, Decision Tree Classifier, Dummy Classifier, Extra Trees Classifier, Gradient Boosting Classifier, K Neighbors Classifier, Light Gradient Boosting Machine, Linear Discriminant Analysis, Logistic Regression, Naive Bayes, Quadratic Discriminant Analysis, Random Forest Classifier, Ridge Classifier, SVM – Linear Kernel, Ada Boost Classifier. To evaluate the performance of each model, we calculated and compared the AUROC with the confidence intervals using the PyCaret package.

## MCCP analysis

Conformal prediction (CP) is a method used to estimate the confidence of predictions for new instances, based on historical data sets. Rather than providing a single-point prediction, CP produces a set of possible outcomes with a validity guarantee: under the assumption that data are exchangeable, the true label lies within the prediction region with probability at least 1 − α, where α is a user-defined error level [16, 21].

A key component of CP is the Nonconformity Measure (NCM), which quantifies how unusual or nonconforming an observation is with respect to the model built from the training data. The NCM is calculated for each individual in both the calibration and test sets and serves as the basis for determining which labels should be included in the prediction region [16]. CP is model-agnostic and can be applied on top of any machine learning algorithm that produces predictive outputs.

In this study, we implemented MCCP using LR as the base model, following the CP framework.

We used the ADVANCE European ancestry population as a training population to build and calibrate the model, while the UKBB WB, AFR and SA populations were used separately as test populations to estimate probabilities and evaluate model performance. The training set was randomly divided into *k* folds (*k* = 5). For each fold, *k* − 1 subsets were used as the training set to fit an LR model using R’s glmnet package, while the remaining subset was used as the calibration set. A prediction region was defined for each individual using the MCCP method, which accounts for class imbalance[15, 17]. Based on the NCM, a prediction region was computed for each test observation.

Depending on the values of the class-specific probabilities *p*1 (case) and *po* (control), and user-defined error level α the prediction can be classified as: case (*p*1 > α *and p*0 ≤ α), control (*p*0 > α *and p*1 ≤ α), uncertain(*p*1 > α *and p*0 > α), or unpredictable (*p*1 < α *and p*0 < α) with a confidence level of 1 − α, credibility corresponds to the probability associated with the most probable class, i.e., the max (*p*1, *p*0)[15, 17]. Coverage refers to the proportion of individuals for whom the model issues a confident prediction (i.e., classified as either case or control), excluding uncertain or unpredictable classifications.

## Logistic Regression – based Stratification by Predicted Probabilities (hereafter referred to as LR-SP)

For the LR-SP model, we selected subsets of individuals based on the predictive probabilities obtained from a LR model fitted using R’s glmnet package. Individuals were classified according to upper and lower thresholds derived from the quantiles of predictive probabilities. The top individuals were those whose predicted probabilities fell in the upper quantiles (e.g., the highest 10%, 5%, or 1%), while the bottom individuals were those in the lower quantiles (e.g., the lowest 10%, 5%, or 1%). This stratification enables targeted evaluation of model performance in groups with extreme predicted outcomes and helps assess the model’s behavior at different levels of prediction confidence.

In the first step, both methods LR-SP and MCCP were applied independently to the three target populations: SA, WB, AFR, using the actual sample sizes specific to each population, without prior adjustment. In the second step, to enable balanced comparisons across populations, stratified random subsampling was applied to the SA and WB datasets, in order to reduce their sample sizes to match the AFR population for each phenotype.

## Results

The baseline characteristics of 17,574 individuals with T2D of self-declared White British origin (WB), 1,145 of genetically determined South-Asian (SA) and 749 of genetically determined African (AFR) origin of the UK Biobank are shown in Table 1. Compared to the two other ethnic groups, WB were slightly older and had diabetes at an older age resulting in a slightly shorter diabetes duration. Their risk factors such as HbA1c levels, blood pressure and BMI were only slightly different. The frequency of individuals with macroalbuminuria was higher in SA and AFR than WB. Stroke to myocardial infarction ratio was close to one in AFR group while the other ethnic groups had a higher frequency of myocardial infarction compared to stroke. While statistical differences exist, their clinical effect sizes are modest.

**Table 1.** Characteristics of participants with T2D of White British (**WB**), South Asians (**SA**), and Africans (AFR) descent from UK Biobank.

| Characteristics | UKBB (n = 19,468) |  |  | P |  |  |
| --- | --- | --- | --- | --- | --- | --- |
|  | WB (n = 17,574) | SA (n = 1,145) | AFR (n = 749) | WB vs SA | WB vs AFR | SA vs AFR |
| Age mean, years (SD) | 60.8 (6.6) | 57.9 (7.5) | 56.9 (8.2) | <0.001 | <0.001 | 0.014 |
| Female, n, (%) | 6,273 (35.7) | 385 (33.6) | 374 (49.9) | 0.156 | <0.001 | <0.001 |
| Diagnosis age mean, years (SD) | 54.6 (8.4) | 50.2 (8.4) | 49.9 (8.9) | <0.001 | <0.001 | 0.272 |
| Diabetes duration mean, years (SD) | 6.2 (6.1) | 7.7 (6.7) | 7 (6.7) | <0.001 | 0.002 | 0.004 |
| BMI mean (kg/m<sup>2</sup>) (SD) | 31.8 (5.8) | 28.9 (5) | 31.7 (6) | <0.001 | 0.215 | <0.001 |
| HbA1c % mean (SD) | 6.9 (1.2) | 7.2 (1.3) | 7.3 (1.7) | <0.001 | <0.001 | 0.717 |
| SBP mean mmHg (SD) | 144 (18) | 141 (18) | 144 (20) | <0.001 | 0.116 | 0.004 |
| DBP mean mmHg (SD) | 82 (10) | 82 (10) | 84 (11) | 0.280 | <0.001 | <0.001 |
| eGFR,ml/min/1.72m<sup>2</sup>,Median(IQR) | 90.7 (79-98) | 95.2 (84-103) | 98.7 (82-111) | <0.001 | <0.001 | <0.001 |
| Low_eGFR | 1,124 (6.7) | 68 (6.3) | 47 (6.6) | 0.611 | 0.904 | 0.810 |
| Macroalbuminuria, n (%) | 305 (4.4) | 34 (8.3) | 27 (8.3) | <0.001 | 0.001 | 0.986 |
| Stroke, n (%) | 1,264 (7.2) | 68 (5.9) | 43 (5.7) | 0.110 | 0.132 | 0.858 |
| MI, n (%) | 2,383 (13.6) | 188 (16.4) | 50 (6.7) | 0.006 | <0.001 | <0.001 |

n : number of patients, yr: Year, SD: Standard deviation, BMI: Body mass index, HbA1c: Glycated hemoglobin, SBP: Systolic blood pressure, DBP: Diastolic blood pressure, UKBB : United Kingdom Biobank, AFR: Africans, SA : South Asians, WB : White British, ADVANCE : Action in Diabetes and Vascular Disease: Preterax and Diamicon Modified Release Controlled Evaluation, MI: myocardial infarction, Low eGFR: estimated glomerular filtration rate based on the CKD-EPI formula (2009) < 60 ml/min/1.72m<sup>2</sup>][22], AFR : Africans, SA : South Asians, WB : Whites British.

We evaluated the potential of various statistical and machine learning methods to predict the risk of stroke, MI and low eGFR in patients with T2D. We used the ADVANCE cohort as the training cohort and the three UK Biobank populations to test the models. The input features were the 10 PRS, the age at the onset of diabetes, its duration, sex and 4 PC of genetic ancestry. The performance metric considered in these analyses was the area under the ROC curve (AUROC). Linear discriminant analysis (LDA), ridge classifier and logistic regression (LR) performed the best, with similar AUROC between the three ethnic groups (Figure 1). The superior performance achieved by these three statistical methods is likely due to the underlying structure of the input data. Linear methods remain the most appropriate for this type of structure. Their better performance can also be attributed to the greater ability of linear models to generalize with smaller datasets, unlike other classification methods that often require larger training cohorts, particularly due to their need for extensive tuning. We therefore continued to use LR to predict the three main diabetes outcomes in the three ethnic groups.

**Figure 1:**
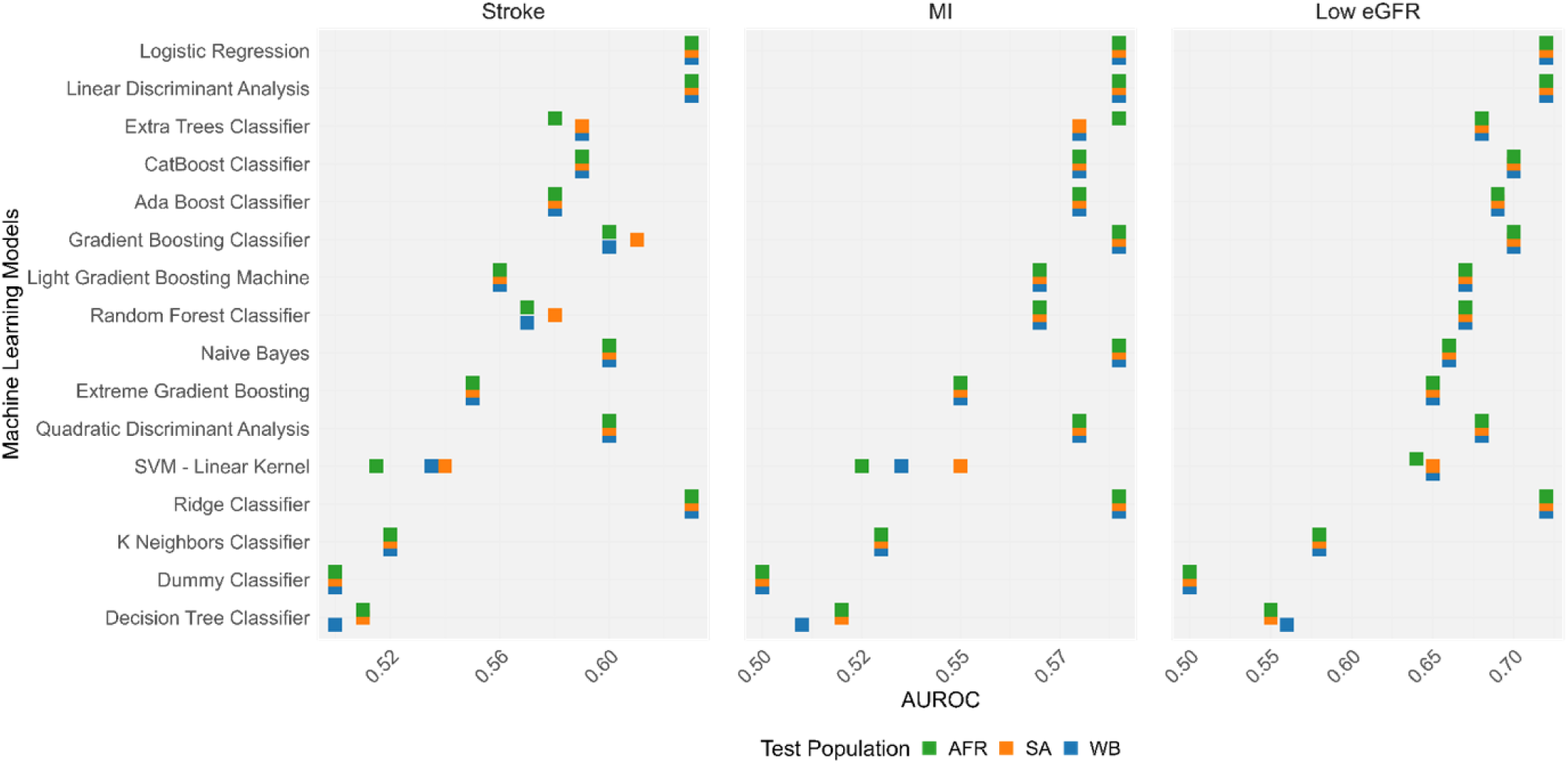
Performance (**AUROC**) of multiple algorithms for diabetes complications across **UKBB** populations using **ADVANCE** as the training Dataset. **MI**: Myocardial infarction, **Low eGFR**: estimated glomerular filtration rate based on the CKD-EPI formula (2009) < 60 ml/min/1.72m²)[22], **AUROC**: Area Under the Receiver Operating Characteristic Curve, **AFR**: Africans, **SA**: South Asians, **WB**: Whites British.

## Performance of MCCP and LR-SP in the prediction of cardiovascular and renal complications of T2D in the three UKBB populations

Having established the most suitable modeling approach based on overall performance, we next assessed two complementary strategies to improve prediction interpretability and confidence estimation across populations: LR-SP and MCCP. At each of the population coverage levels, the AUROC obtained with MCCP were consistently higher than those with LR-SP for the three complications at error rates α ranging from 0.05 to 0.4 (α representing the tolerated probability that the true label is *not* in the predicted region). This indicates that MCCP improves predictive accuracy by selectively retaining cases where the model has high confidence (Fig.2, page 29).

**Fig. 2.**
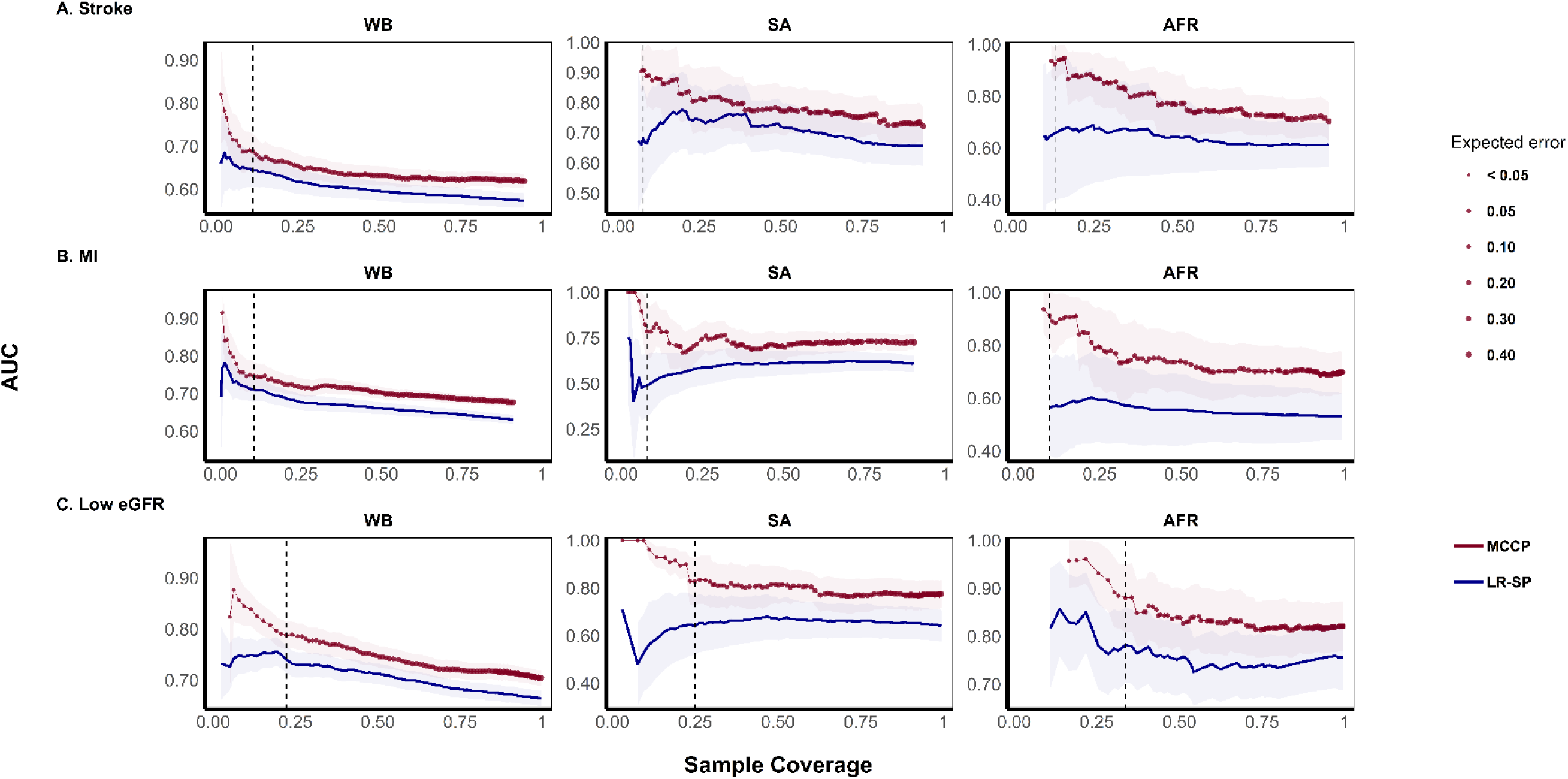
Comparison of AUROC performance between MCCP and the LR-SP method for predicting cardiovascular and renal complications of diabetes (stroke, MI, and low eGFR). Models included 10 weighted polygenic risk scores (wPRS), sex, age at diagnosis of type 2 diabetes (T2D), diabetes duration, and the first four principal components (PC1–PC4) of genetic ancestry. The x-axis represents sample coverage. For LR-SP, this corresponds to the proportion of individuals with predicted probabilities in the extreme tails of the distribution (e.g., top and bottom x%) classified as cases or controls. For MCCP, coverage refers to the proportion of individuals for which a prediction is made, with point size indicating the expected error (up to 0.40). Dashed vertical lines denote an expected error of 0.05 for MCCP. Solid lines and shaded areas indicate median AUROC and 95% confidence intervals, respectively.

In WB population (Table 2A, page 27; Fig. 2, page 29; Suppl. Figs. S1–S3), at an error rate of α = 0.05, MCCP predicted 12.3% of subjects (AUROC = 0.70; 95% CI: 0.64–0.72) for stroke, 10.4% for MI (AUROC = 0.75; 95% CI: 0.71–0.79), and 22.3% for low eGFR (AUROC = 0.79; 95% CI: 0.76–0.81). Compared to LR-SP that selects the 5% of top high– and bottom low-risk individuals according to the predicted probability quantiles (ensuring 10% coverage), MCCP showed an improvement in AUROC by 0.05 for stroke, 0.04 for MI and 0.04 for low eGFR. Low eGFR was predicted with the highest AUROC and coverage with MCCP in the WB population. MCCP did not modify the positive predictive values (PPV) while the negative predictive values (NPV) were high with or without MCCP, reaching 0.98 for low eGFR, 0.97 for stroke and 0.95 for MI, suggesting the model is very good at ruling out individuals who won’t have the outcome. MCCP improved case detection (recall), with a gain of 0.04 for stroke (0.83 vs. 0.79) thus reducing the number of false negatives. However, for MI and low eGFR, the recall was lower than with LR-SP (0.66 vs. 0.84) and (0.82 vs 0.94) suggesting that MCCP favors overall discrimination at the expense of sensitivity.

**Table 2.** Comparison of the performance of MCCP (at an expected error of 0.05) and the LR-SP method (top and bottom 5% of predicted probabilities). Models were trained on individuals of European descent (EUR) from the ADVANCE study, using 10 polygenic risk scores (PRS), sex, age at type 2 diabetes (T2D) diagnosis, duration of T2D, and the first four genetic principal components (PC1–PC4) as covariates. The test populations from the UK Biobank were analyzed separately: (A) White British (WB), (B) South Asian (SA), and (C) African (AFR). MI: Myocardial infarction; Low eGFR: estimated glomerular filtration rate < 60 ml/min/1.73 m² (CKD-EPI 2009).

| Disease | Method | Coverage (%) | AUROC (95% CI) | PPV (95% CI) | NPV (95% CI) | RECALL (95% CI) |
| --- | --- | --- | --- | --- | --- | --- |
| Stroke | MCCP | 12.3 | 0.70 (0.64-0.72) | 0.12 (0.11-0.14) | 0.97 (0.96-0.98) | 0.83 (0.65-0.88) |
|  | Classical | 10 | 0.65 (0.61-0.69) | 0.12 (0.11-0.14) | 0.97 (0.96-0.98) | 0.79 (0.70-0.86) |
| MI | MCCP | 10.4 | 0.75 (0.71-0.79) | 0.22 (0.17-0.26) | 0.95 (0.94-0.97) | 0.66 (0.56-0.79) |
|  | Classical | 10 | 0.71 (0.68-0.74) | 0.22 (0.21-0.24) | 0.96 (0.95-0.97) | 0.84 (0.78-0.88) |
| Low eGFR | MCCP | 22.3 | 0.79 (0.76-0.81) | 0.13 (0.12-0.17) | 0.98 (0.98-0.99) | 0.82 (0.68-0.89) |
|  | Classical | 10 | 0.75 (0.71- 0.78) | 0.15 (0.14-0.17) | 0.99 (0.98-1.00) | 0.94 (0.86-0.98) |

B) SA
| Disease | Method | Coverage (%) | AUROC (95% CI) | PPV (95% CI) | NPV (95% CI) | RECALL (95% CI) |
| --- | --- | --- | --- | --- | --- | --- |
| Stroke | MCCP | 8.1 | 0.91 (0.84-0.97) | 0.35 (0.26-0.58) | 0.99 (0.99-1.00) | 0.99 (0.88-1.00) |
|  | Classical | 10 | 0.71 (0.56-0.86) | 0.20 (0.16-0.27) | 0.99 (0.96-1.00) | 0.99 (0.96-1.00) |
| MI | MCCP | 9.2 | 0.78 (0.64-0.93) | 0.30 (0.20-0.71) | 0.97 (0.93-1.00) | 0.83 (0.50-1.00) |
|  | Classical | 10 | 0.50 (0.33-0.67) | 0.19 (0.13-0.30) | 0.92 (0.87-1.00) | 0.64 (0.21-1.00) |
| Low eGFR | MCCP | 24.9 | 0.83 (0.73-0.93) | 0.10 (0.08-0.45) | 0.99 (0.98-1.00) | 0.99 (0.64-1.00) |
|  | Classical | 10 | 0.56 (0.38-0.74) | 0.13 (0.07-0.19) | 0.97 (0.95-1.00) | 0.75 (0.50-1.00) |

C) AFR
| Disease | Method | Coverage (%) | AUROC (95%CI) | PPV (95%CI) | NPV (95%CI) | RECALL (95%CI) |
| --- | --- | --- | --- | --- | --- | --- |
| Stroke | MCCP | 12.1 | 0.94 (0.88-1.00) | 0.37 (0.27-0.67) | 0.99 (0.99-1.00) | 0.99 (0.99-1.00) |
|  | Classical | 10 | 0.64 (0.36-0.92) | 0.36 (0.12-1.00) | 0.95 (0.92-1.00) | 0.57 (0.14-1.00) |
| MI | MCCP | 9.6 | 0.91 (0.81-1.00) | 0.33 (0.21-1.00) | 0.99 (0.97-1.00) | 0.99 (0.71-1.00) |
|  | Classical | 10 | 0.57 (0.38-0.76) | 0.20 (0.15-0.36) | 0.95 (0.89-1.00) | 0.80 (0.40-1.00) |
| Low eGFR | MCCP | 33.6 | 0.88 (0.81-0.95) | 0.17 (0.14-0.44) | 0.99 (0.99-1.00) | 0.99(0.79-1.00) |
|  | Classical | 10 | 0.82 (0.69-0.94) | 0.33 (0.18-0.60) | 0.99 (0.96-1.00) | 0.88 (0.75-1.00) |

In the SA population (Table 2B, page 27; Fig. 2, page 29, Supplementary Fig. 1–3), MCCP predicted at an error rate α = 0.05, 8.1% of individuals for stroke, 9.2% for MI, and 24.9% for low eGFR with AUROC of 0.91 (95% CI: 0.84–0.97), 0.78 (95% CI: 0.64–0.93) and 0.83 (95% CI: 0.73–0.93), respectively. Compared to LR-SP, MCCP showed a marked improvement in AUROC by 0.20 for stroke (0.91 vs. 0.71), 0.28 for MI (0.78 vs. 0.50) and 0.27 for low eGFR (0.83 vs. 0.56) in this population. As in WB, low eGFR had the highest coverage of the population with a predicted error rate of 0.05. PPV was also higher with MCCP than with LR-SP (0.35 vs. 0.20) for stroke and 0.30 vs. 0.19 for MI. MCCP did not improve low eGFR AUROC (0.10 vs. 0.13). As expected, NPV was very high with or without MCCP, reaching 0.99 for stroke, 0.97 for MI and 0.99 for low eGFR. Most importantly, MCCP improved markedly case detection in SA for all three complications (0.99 for stroke, 0.83 for MI and 0.99 for low EGFR).

In the AFR population (Table 2C, page 28; Fig. 2, page 29, Supplementary Fig. 1–3) at an error rate α = 0.05, MCCP predicted 12.1% of individuals for stroke, 9.6% for MI and 33.6% for low eGFR with AUROC of 0.94 (95% CI: 0.88–1.00), 0.91 (95% CI: 0.81–1.00) and 0.88 (95% CI: 0.81–0.95), respectively. Compared to LR-SP, MCCP significantly improved AUROC, PPV and recall in the AFR population.

Theses analyses showed that MCCP improved the AUROC in the three populations and for the three diabetes complications. The improvement was more important in AFR and SA than in WB. Low eGFR consistently had the highest coverage at the error rate of α = 0.05 in the three populations. In addition to improving discrimination, MCCP also enhanced recall by increasing sensitivity and reducing false negatives within the high-confidence prediction subset.

## Performance of MCCP and LR-SP after sample size adjustments

We uniformized population sizes by applying stratified random subsampling based on case–control status, reducing the WB and SA populations to match the AFR sample size while preserving class balance for each phenotype. The distributions of key covariates (age at diagnosis, diabetes duration, sex, and principal components, PC1–PC4) were maintained between the original and downsampled datasets. Statistical tests (Kolmogorov–Smirnov[23] for continuous variables and Fisher’s exact test[24] for categorical variables) revealed no significant differences, confirming the representativeness of the downsampled samples (Supplementary Table 1-2, Supplementary Fig. 4–9). This adjustment was intended to determine whether the observed differences between WB, SA and AFR populations were due to differences in sample size or methodological biases. Apart from the NPV that was nearly at its maximum in the three populations as in the previous analyses, reducing the sample size of WB and SA led to increase in coverage, AUROC, PPV and recall measured by the MCCP for the three complications. These parameters were also increased with LR-SP that had a fixed coverage at 10%. At same sample size (Table 3, page 30,31; Fig.3, page 32; Supplementary Fig. 10–12)., coverage for stroke with MCCP, at an error rate α = 0.05, remained the highest in WB (18.7%), followed by SA (12.1%) and AFR (12.1%) with AUROC reaching 0.99 in WB, 0.96 in SA and 0.94 in AFR. Coverage remained also the highest for MI after sample size reduction among WBs (14.3%), followed by SAs (9.7%) and AFRs (9.6%) with AUROC reaching 0.96 in WBs, 0.87 in SA and 0.91 in AFR. Low eGFR had a distinct profile from stroke and MI complications. Coverage was the highest among AFRs (33.6%), followed by SAs (27.5%) and WBs (23%) while AUROC remained the highest in WB (0.92), followed by AFR (0.88) and SA (0.85).

**Fig. 3.**
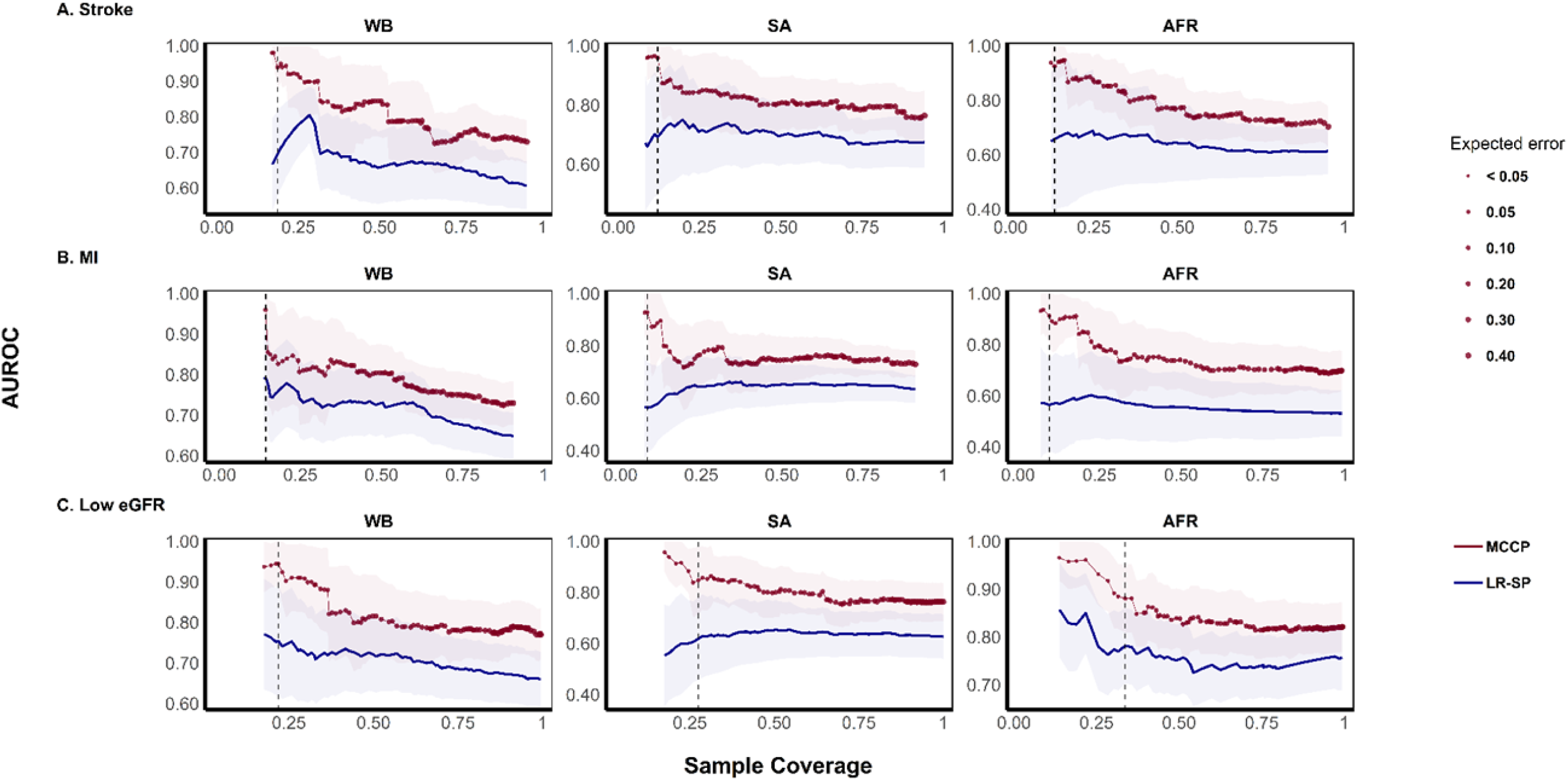
Comparison of MCCP and LR-SP performance using test sets of equal size. WB and SA populations were downsampled using stratified random subsampling based on case–control status to match the sample size of the AFR group. All other parameters and inputs are as described in Fig. 2.

**Table 3.** Comparison of MCCP (expected error = 0.05) and LR-SP (top and bottom 5% of predicted probabilities) after harmonizing test set sizes. WB and SA samples were downsampled using stratified random subsampling (based on case–control status) to match the sample size of the AFR population. Model inputs and training procedures were identical to Table 2. MI: Myocardial infarction; Low eGFR: estimated glomerular filtration rate < 60 ml/min/1.73 m² (CKD-EPI 2009).

| Disease | Method | Coverage (%) | AUROC (95% CI) | PPV (95% CI) | NPV (95% CI) | RECALL (95% CI) |
| --- | --- | --- | --- | --- | --- | --- |
| Stroke | MCCP | 18.7 | 0.99 (0.98-1.00) | 0.40 (0.26-1.00) | 0.99 (0.98-1) | 0.99 (0.98-1.00) |
|  | Classical | 10 | 0.80 (0.52-1.00) | 0.07 (0.05-0.40) | 0.99 (0.98-1) | 0.99 (0.98-1.00) |
| MI | MCCP | 14.3 | 0.96 (0.91-1.00) | 0.39 (0.27-1.00) | 0.99 (0.99-1) | 0.99 (0.89-1.00) |
|  | Classical | 10 | 0.79 (0.67-0.91) | 0.35 (0.21-0.56) | 0.96 (0.92-1) | 0.79 (0.57-1.00) |
| Low eGFR | MCCP | 23 | 0.92 (0.85-1.00) | 0.32 (0.14-0.80) | 0.99 (0.98-1) | 0.90 (0.70-1.00) |
|  | Classical | 10 | 0.72 (0.57-0.87) | 0.23 (0.19-0.43) | 0.99 (0.96-1) | 0.99 (0.75-1.00) |

B) SA
| Disease | Method | Coverage (%) | AUROC (95% CI) | PPV (95% CI) | NPV (95% CI) | RECALL (95% CI) |
| --- | --- | --- | --- | --- | --- | --- |
| Stroke | MCCP | 12.1 | 0.96 (0.87-1.00) | 0.83 (0.25-1.00) | 0.98 (0.98-1.00) | 0.98 (0.83 -1.00) |
|  | Classical | 10 | 0.67 (0.46-0.87) | 0.20 (0.12-0.50) | 0.98 (0.93-1.00) | 0.86 (0.43-1.00) |
| MI | MCCP | 9.7 | 0.87 (0.68-1.00) | 0.50 (0.3-1.00) | 0.98 (0.95-1.00) | 0.86 (0.57-1.00) |
|  | Classical | 10 | 0.57 (0.41-0.73) | 0.23 (0.18-0.35) | 0.95 (0.87-1.00) | 0.83 (0.42-1.00) |
| Low eGFR | MCCP | 27.5 | 0.85 (0.72-0.98) | 0.16 (0.08-0.64) | 0.99 (0.98-1.00) | 0.88 (0.62-1.00) |
|  | Classical | 10 | 0.60 (0.37-0.83) | 0.10 (0.08-0.29) | 0.99 (0.97-1.00) | 0.10 (0.50 -1.00) |

C) AFR
| Disease | Method | Coverage (%) | AUROC (95% CI) | PPV (95% CI) | NPV (95% CI) | RECALL (95% CI) |
| --- | --- | --- | --- | --- | --- | --- |
| Stroke | MCCP | 12.1 | 0.94 (0.88-1.00) | 0.37 (0.27-0.67) | 0.99 (0.99-1.00) | 0.99 (0.99-1.00) |
|  | Classical | 10 | 0.64 (0.36-0.92) | 0.36 (0.12-1.00) | 0.95 (0.92-1.00) | 0.57 (0.14-1.00) |
| MI | MCCP | 9.6 | 0.91 (0.81-1.00) | 0.33 (0.21-1.00) | 0.99 (0.97-1.00) | 0.99 (0.71-1.00) |
|  | Classical | 10 | 0.57 (0.38-0.76) | 0.20 (0.15-0.36) | 0.95 (0.89-1.00) | 0.80 (0.40-1.00) |
| Low eGFR | MCCP | 33.6 | 0.88 (0.81-0.95) | 0.17 (0.14-0.44) | 0.99 (0.99-1.00) | 0.99(0.79-1.00) |
|  | Classical | 10 | 0.82 (0.69-0.94) | 0.33 (0.18-0.60) | 0.99 (0.96-1.00) | 0.88 (0.75-1.00) |

These results indicate that the initial performance gap was due to significant differences in population size. Once adjusted to the same size, the higher predictive performance observed in the WB could be attributed to its greater genetic proximity to the reference population used for training.

## Discussion

The under representation of populations of non-European ancestry in genetic and genomic studies has major implications for health equity. It limits the understanding of the genetic factors involved in diseases and results in prediction tools being less accurate or applicable in these populations, reinforcing disparities in access to predictive medicine. Polygenic risk scores (PRS), largely developed using European cohorts, tend to perform poorly in more genetically diverse populations, especially among individual of Africans ancestry. This disparity may be explained by structural differences, including linkage disequilibrium (LD) patterns, allele frequency distribution, and population-specific environmental influences [25–29].

MCCP is a machine learning confidence estimation framework that offers an interesting avenue for the clinical application of PRS, in particular to improve the reliability and interpretability of genetic risk estimates, particularly in imbalanced or heterogeneous populations [15, 17]. Unlike conventional approaches based on arbitrary thresholds, MCCP offers an individualized estimate of risk while ensuring explicit control of the error rate. Its main advantage is its optimized calibration, which ensures that the proportion of erroneous predictions remains within the specified error rate [15, 17].

In this work, we compared multiple machine learning methods and selected logistic regression (LR) based on AUROC performance. We then compared two LR-based approaches: LR-based predicted probability stratification (LR-SP), and Mondrian Cross-Conformal Prediction (MCCP) using LR as the underlying algorithm. These analyses aimed to assess the transferability of our multiPRS, developed in individuals of European descent (EUR) from the ADVANCE clinical trial to three target populations from the UK Biobank: White British (WB), South Asian (SA), and African (AFR)

It should be noted that the study of the three populations living in the United Kingdom, thus sharing a similar environment, reduces broad environmental variations between different countries. However, differences in social determinants of health, lifestyle, and healthcare access may still exist and influence both risk and prediction performance.

Our results indicate that the use of MCCP consistently improves the performance of the multiPRS in terms of AUROC, surpassing LR-SP in the three populations studied. Expectedly, with equal sample size, the highest AUROC for stroke and MI were observed in participants of WB, likely reflecting their greater genetic proximity to the EUR population used for the training of the ADVANCE model, which supports the idea that PRS transferability tends to be greater between genetically similar populations, a phenomenon well documented in the literature [27, 30].

However, unlike stroke and MI, where a higher coverage in WBs was accompanied by a stronger AUROC, the relationship between coverage and performance for low eGFR appears to follow a different trend. This suggests that, beyond genetic proximity, other factors may play a role in PRS performance for predicting low eGFR, possibly related to its distinct genetic and pathophysiological architecture, as well as to a more subtle correlation between eGFR and cardiovascular diseases. This has been reported in a recent study that highlighted an inverse-non-linear correlation that persists between eGFR and cardiovascular disease risk but becomes non-significant at certain levels of eGFR[31].

Additionally, the method used to estimate eGFR may have contributed to this discrepancy. In this study, eGFR was calculated using the CKD-EPI equation including the correction factor for individuals of African ancestry [22]. This correction, historically justified by presumed differences in average muscle mass, systematically increases the estimated eGFR in this population [22], potentially leading to the misclassification of true CKD cases as non-cases. It has been shown that removing the ethnicity-specific coefficient would reclassify over one-third of African American patients into a more advanced CKD stage, with significant clinical consequences [32, 33].

Other studies have reported that genetically determined African ancestry is associated with creatinine and eGFR levels, independently of self-reported ethnicity, and that APOL1 variants are strongly associated with CKD risk [34]. More recently, new equations combining creatinine and cystatin C, without incorporating ethnicity, have been proposed to improve equity in kidney function assessment[35]. Moreover, ancestry-specific loci associated with eGFR have been identified through admixture mapping, highlighting the influence of local ancestral genomic segments on this phenotype [36]. Altogether, these findings indicate that eGFR should not be considered a uniform clinical trait across populations, and that its estimation shaped by methodological choices and population specific genetic factors can significantly impact PRS performance in multi-ancestry analyses.

However, the improvements observed for positive predictive value (PPV) and recall (sensitivity) vary across phenotypes and populations studied. In some cases, PPV and recall remain comparable to those of LR-SP, while in others, MCCP offers a significant gain. PPV remained moderate in all populations studied. This is because PPV values are constrained by event rarity in the target population, a phenomenon that is well documented in epidemiology and predictive medicine [37, 38].

Our results obtained with MCCP in SA and AFR provide a way that our multiPRS, although developed from European populations, could still provide meaningful risk stratification in more genetically diverse groups. These findings suggest that, when coupled with advanced approaches like MCCP, PRS could be adapted beyond their original population to support cautious application of PRS in genetically heterogeneous settings, even when full re-development of the score is not feasible.

A major advantage of the MCCP method is its ability to evaluate confidence intervals for each individual prediction. These personalized estimates can allow practitioners to make clinical decisions based on a specific expected error (α). Thus, the MCCP offers a valuable tool to guide medical judgment, providing not only predictions, but also a measure of their reliability for each patient [17].

While these results are promising, there are several limitations to consider. First, the SA and AFR sample sizes remain relatively small. This highlights the importance of increasing the recruitment of participants from underrepresented populations. This need is all the more pressing as recruitment in these groups has stagnated, or even declined, in recent years, with the proportion of genetic data generated for individuals of EUR ancestry increasing from 86.3% in 2021[39] to 94.48.% in 2024[40].

In addition, the large intra-African heterogeneity complicates the transferability of PRSs, as genetic and environmental differences within the continent influence the performance of the models. For example, one study [39] showed that the PRS developed in African Americans are not always applicable to African populations, due to differences in genetic architecture, allele frequencies, and the fact that African Americans represent only a subset of West African ancestry with admixture Specifically, genetic and environmental gaps between the Ugandan (East Africa) and South African cohorts contributed to the low transferability of genetic risk scores (GRS) derived from African Americans, highlighting the need to adapt PRS according to the specific genetic and environmental context of each population [39].

## Conclusion

Our results show that our multiPRS developed from populations of EUR origin could be applied to populations of SA and AFR origin, especially when coupled with advanced methods such as Mondrian Cross-Conformal Prediction (MCCP). By enabling better calibration of predictions and explicit control of the error rate, MCCP significantly improves the accuracy and robustness of predictive models, while maintaining competitive performance in all populations studied.

Although WB participants are the best performers (at same sample size), the results obtained in SA and AFR are encouraging, highlighting the ability of the multiPRS to be adapted beyond their initial training population. These observations reinforce the idea that PRSs developed in EUR populations may be cautiously extended to genetically diverse groups when combined with approaches that enable individualized confidence estimation and risk stratification.

In future work, we plan to extend our analysis to larger and more diverse cohorts, including other underrepresented ancestral groups such as Latino and admixed populations, in order to improve the robustness and generalizability of our multiPRS. We also aim to evaluate the MCCP method in a clinical setting, by assessing whether its ability to identify not only cases and controls, but also uncertain or unpredictable situations, can enhance risk stratification and help better target patients requiring more intensive follow-up.

## Data Availability

All data produced in the present study are available upon reasonable request to the authors

## Acknowledgments

This work was supported by grants from Genome Quebec to JT and from the GAPP program of Genome Canada to PH and JT and by OPTI-THERA inc. The analysis was conducted using the UK Biobank Resource under projects 49731 and 59642.

## Author contributions

JT, PH and RA conceived, designed and supervised the whole project. EK and RA performed the analyses. CH contributed to data extraction. JT, RA and EK drafted the manuscript. JH contributed to responsible reporting related to diversity, equity, and population representation in statistical genetics. PH, MS and JH revised the manuscript. All authors have approved the final version of the manuscript.

## Nonstandard Abbreviations and Acronyms

ADVANCE: Action in Diabetes and Vascular Disease: Preterax and Diamicron Modified Release Controlled Evaluation
AFR: African ancestry (UK Biobank)
CP: Conformal Prediction
LR-SP: Logistic Regression–based Stratification by Predicted Probabilities
MCCP: Mondrian Cross-Conformal Prediction
NCM: Nonconformity Measure
MultiPRS: Multi-Polygenic Risk Score
SA: South Asian ancestry (UK Biobank)
WB: White British ancestry (UK Biobank)
wPRS: Weighted Polygenic Risk Score

## Supplementary Information

**Supplementary Fig. 1.**
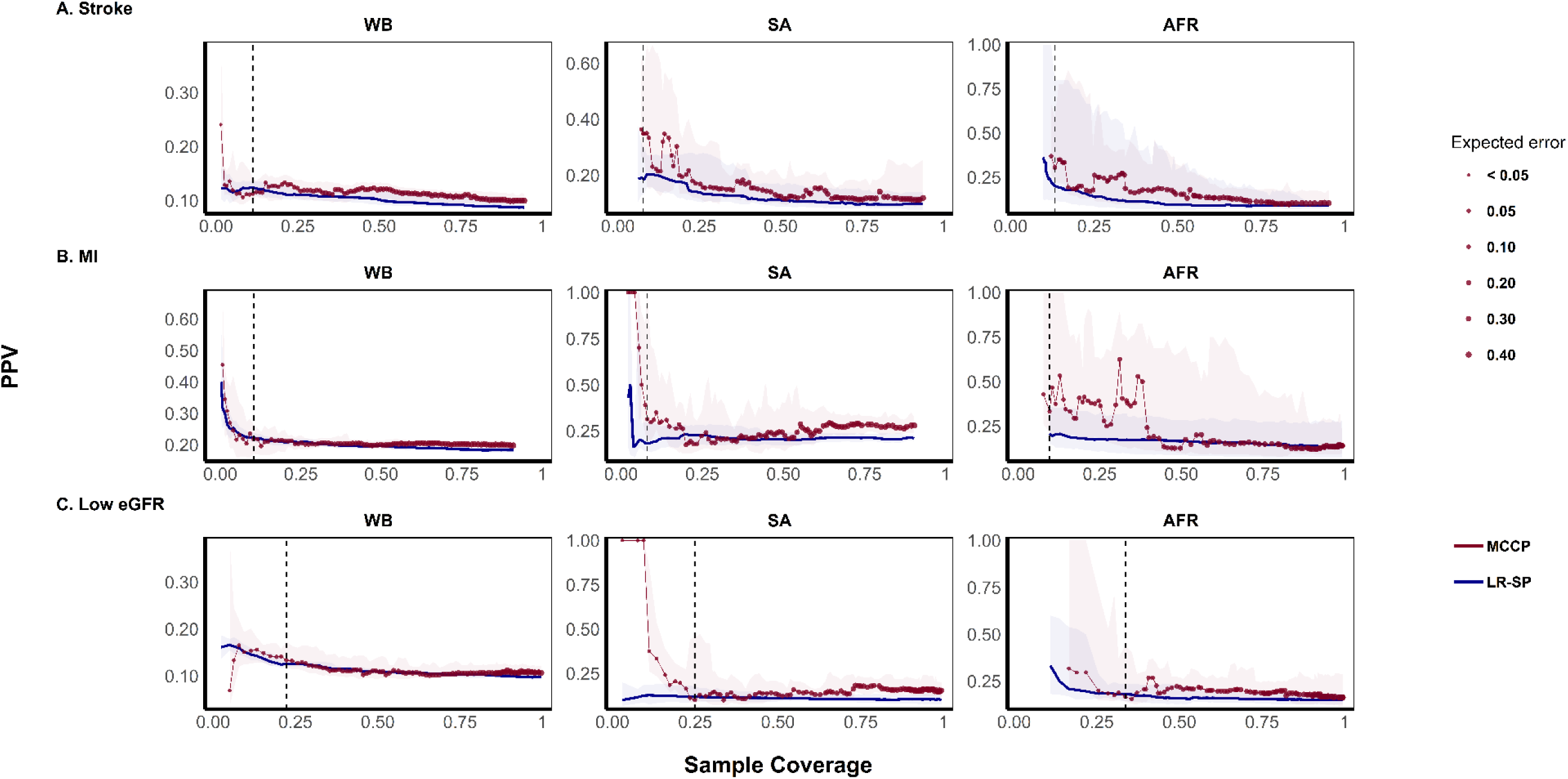
Comparison of PPV between MCCP and the LR-SP method for predicting cardiovascular and renal complications of diabetes (stroke, MI, and low eGFR). Models included 10 weighted polygenic risk scores (wPRS), sex, age at diagnosis of type 2 diabetes (T2D), diabetes duration, and the first four principal components (PC1–PC4) of genetic ancestry. The x-axis represents sample coverage. For LR-SP, this corresponds to the proportion of individuals with predicted probabilities in the extreme tails of the distribution (e.g., top and bottom x%) classified as cases or controls. For MCCP, coverage refers to the proportion of individuals for which a prediction is made, with point size indicating the expected error (up to 0.40). Dashed vertical lines denote an expected error of 0.05 for MCCP. Solid lines and shaded areas indicate median PPV and 95% confidence intervals, respectively.

**Supplementary Fig. 2.**
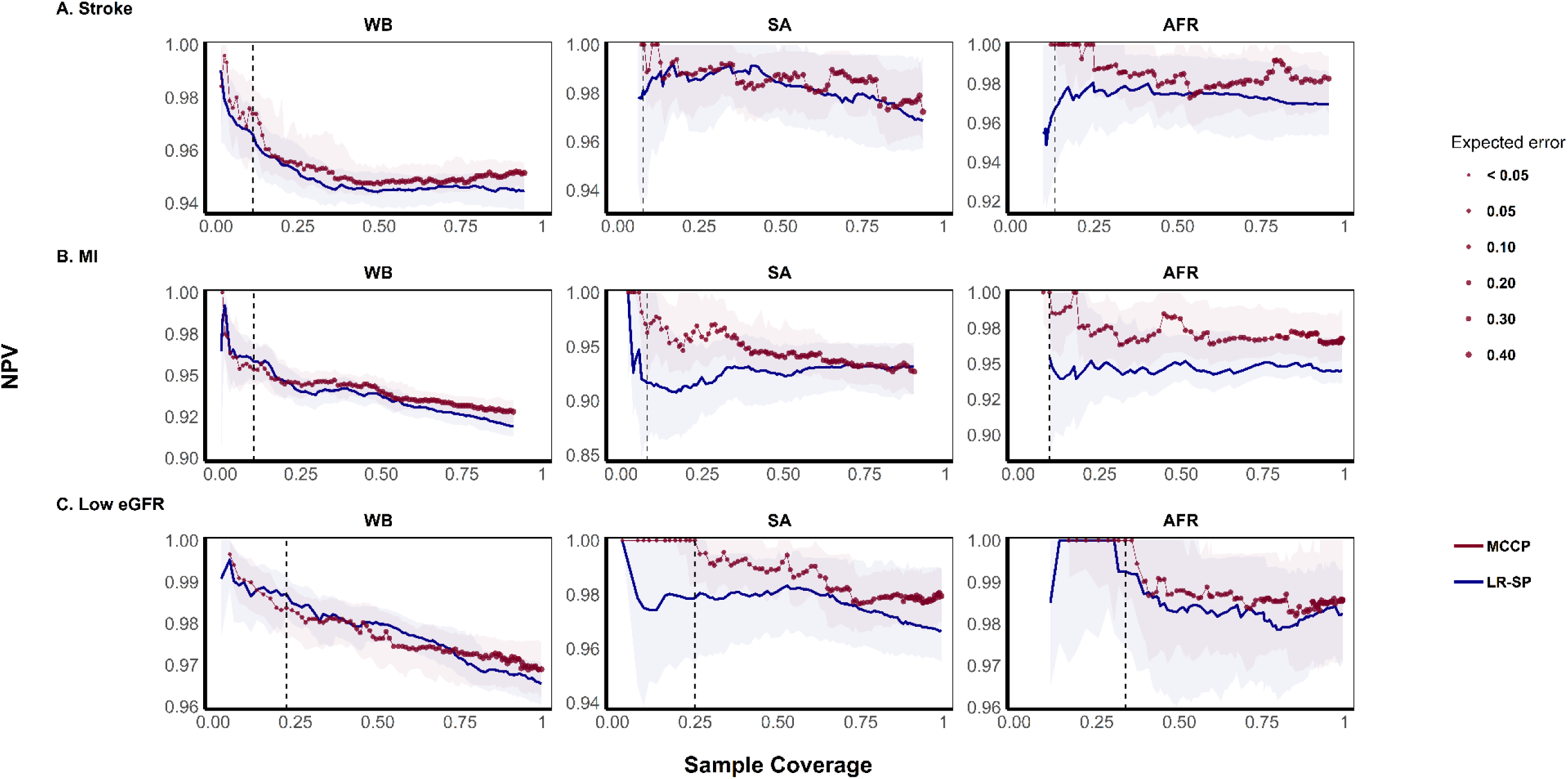
Comparison of NPV between MCCP and the LR-SP method for predicting cardiovascular and renal complications of diabetes (stroke, MI, and low eGFR). Models included 10 weighted polygenic risk scores (wPRS), sex, age at diagnosis of type 2 diabetes (T2D), diabetes duration, and the first four principal components (PC1–PC4) of genetic ancestry. The x-axis represents sample coverage. For LR-SP, this corresponds to the proportion of individuals with predicted probabilities in the extreme tails of the distribution (e.g., top and bottom x%) classified as cases or controls. For MCCP, coverage refers to the proportion of individuals for which a prediction is made, with point size indicating the expected error (up to 0.40). Dashed vertical lines denote an expected error of 0.05 for MCCP. Solid lines and shaded areas indicate median NPV and 95% confidence intervals, respectively.

**Supplementary Fig. 3.**
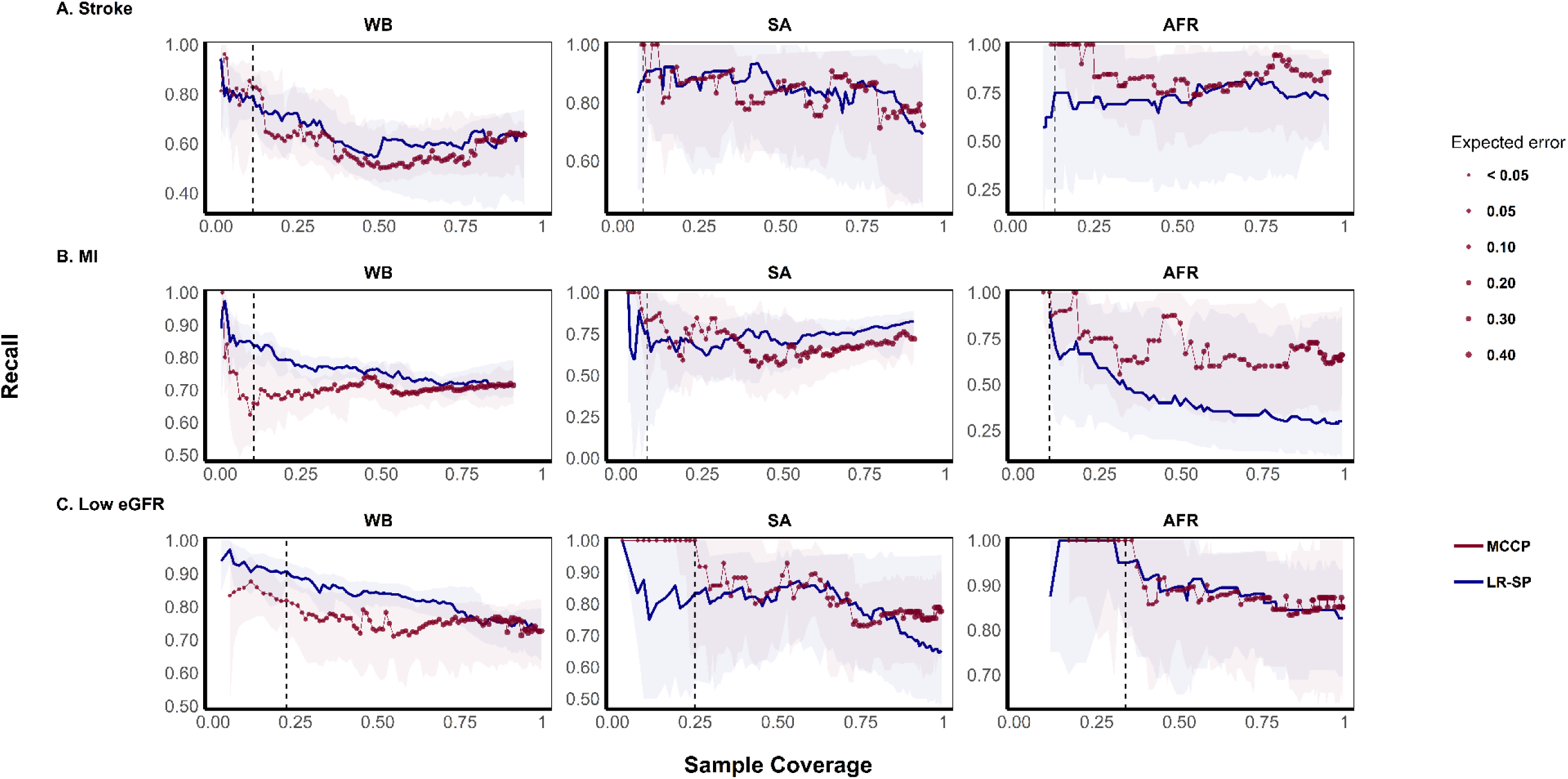
Comparison of Recall between MCCP and the LR-SP method for predicting cardiovascular and renal complications of diabetes (stroke, MI, and low eGFR). Models included 10 weighted polygenic risk scores (wPRS), sex, age at diagnosis of type 2 diabetes (T2D), diabetes duration, and the first four principal components (PC1–PC4) of genetic ancestry. The x-axis represents sample coverage. For LR-SP, this corresponds to the proportion of individuals with predicted probabilities in the extreme tails of the distribution (e.g., top and bottom x%) classified as cases or controls. For MCCP, coverage refers to the proportion of individuals for which a prediction is made, with point size indicating the expected error (up to 0.40). Dashed vertical lines denote an expected error of 0.05 for MCCP. Solid lines and shaded areas indicate median RECALL and 95% confidence intervals, respectively.

**Supplementary Table 1.** Statistical comparison of covariate distributions between the original and downsampled datasets, by phenotype (stroke, myocardial Infarction [MI], and low estimated Glomerular Filtration Rate [Low eGFR]) in the White British (WB) population. Continuous covariates were compared using the Kolmogorov–Smirnov (KS) test, and binary variables using Fisher’s exact test. P-values greater than 0.05 indicate no significant difference in distribution, suggesting that the covariate structure was preserved following downsampling.

| Phenotype | Dataset size (original / downsampled) | Variable | Variable Type | Test | Statistic | P |
| --- | --- | --- | --- | --- | --- | --- |
| **Stroke** | 17,574 / 749 | Stroke (label) | Binary | Fisher | - | 1 |
|  |  | Sex |  |  | - | 0.756 |
|  |  | Diagnosis age | Numeric | KS | 0.021 | 0.914 |
|  |  | Diabetes duration |  |  | 0.02 | 0.934 |
|  |  | PC1 |  |  | 0.025 | 0.774 |
|  |  | PC2 |  |  | 0.019 | 0.96 |
|  |  | PC3 |  |  | 0.039 | 0.218 |
|  |  | PC4 |  |  | 0.031 | 0.487 |
| **MI** | 17,574 / 749 | MI (label) | Binary | Fisher | - | 0.744 |
|  |  | Sex |  |  | - | 0.756 |
|  |  | Diagnosis age | Numeric | KS | 0.021 | 0.914 |
|  |  | Diabetes duration |  |  | 0.02 | 0.934 |
|  |  | PC1 |  |  | 0.025 | 0.774 |
|  |  | PC2 |  |  | 0.019 | 0.96 |
|  |  | PC3 |  |  | 0.039 | 0.218 |
|  |  | PC4 |  |  | 0.031 | 0.487 |
| **Low eGFR** | 16,773 / 713 | Low eGFR (label) | Binary | Fisher | - | 0.189 |
|  |  | Sex |  |  | - | 0.472 |
|  |  | Diagnosis age | Numeric | KS | 0.048 | 0.088 |
|  |  | Diabetes duration |  |  | 0.041 | 0.203 |
|  |  | PC1 |  |  | 0.038 | 0.269 |
|  |  | PC2 |  |  | 0.027 | 0.702 |
|  |  | PC3 |  |  | 0.029 | 0.618 |
|  |  | PC4 |  |  | 0.0370 | 0.293 |

**Supplementary Table 2.** Statistical comparison of covariate distributions between the original and downsampled datasets, by phenotype (stroke, myocardial Infarction [MI], and low estimated Glomerular Filtration Rate [Low eGFR]) in the South Asian (SA) population. Continuous covariates were compared using the Kolmogorov–Smirnov (KS) test, and binary variables using Fisher’s exact test. P-values greater than 0.05 indicate no significant difference in distribution, suggesting that the covariate structure was preserved following downsampling.

| Phenotype | Dataset size (original / downsampled) | Variable | Variable Type | Test | Statistic | P |
| --- | --- | --- | --- | --- | --- | --- |
| **Stroke** | 1,145 / 749 | Stroke (label) | Binary | Fisher | - | 0.688 |
|  |  | Sex |  |  | - | 0.586 |
|  |  | Diagnosis age | Numeric | KS | 0.022 | 0.979 |
|  |  | Diabetes duration |  |  | 0.024 | 0.959 |
|  |  | PC1 |  |  | 0.019 | 0.998 |
|  |  | PC2 |  |  | 0.019 | 0.997 |
|  |  | PC3 |  |  | 0.018 | 0.999 |
|  |  | PC4 |  |  | 0.018 | 0.998 |
| **MI** | 1,144 / 749 | MI (label) | Binary | Fisher | - | 0.801 |
|  |  | Sex |  |  | - | 0.921 |
|  |  | Diagnosis age | Numeric | KS | 0.023 | 0.969 |
|  |  | Diabetes duration |  |  | 0.025 | 0.941 |
|  |  | PC1 |  |  | 0.019 | 0.998 |
|  |  | PC2 |  |  | 0.019 | 0.997 |
|  |  | PC3 |  |  | 0.018 | 0.999 |
|  |  | PC4 |  |  | 0.018 | 0.998 |
| **Low eGFR** | 1,080 / 713 | Low eGFR (label) | Binary | Fisher | - | 0.84 |
|  |  | Sex |  |  | - | 0.758 |
|  |  | Diagnosis age | Numeric | KS | 0.013 | 1 |
|  |  | Diabetes duration |  |  | 0.013 | 1 |
|  |  | PC1 |  |  | 0.017 | 1 |
|  |  | PC2 |  |  | 0.016 | 1 |
|  |  | PC3 |  |  | 0.012 | 1 |
|  |  | PC4 |  |  | 0.014 | 1 |

**Supplementary Fig. 4.**
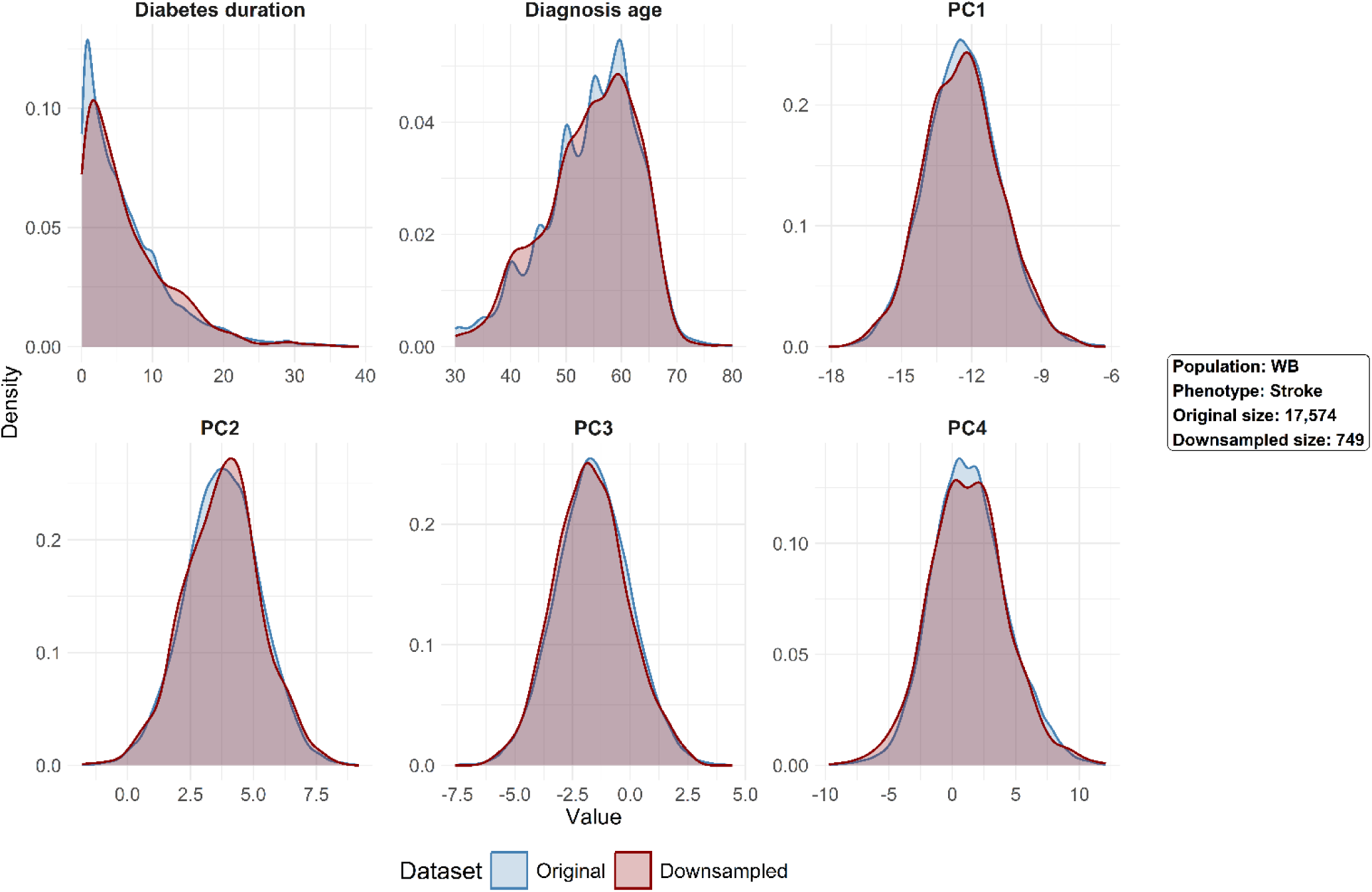
Comparative density plots of continuous covariates used in the predictive models in the White British (**WB**) population for **Stroke**. Distributions are shown for both the original sample and the downsampled dataset. Covariates include age at diabetes diagnosis, diabetes duration, and the first four principal components (**PC1** to **PC4**). Initial distributions were largely preserved after downsampling, indicating good retention of the underlying data structure.

**Supplementary Fig. 5.**
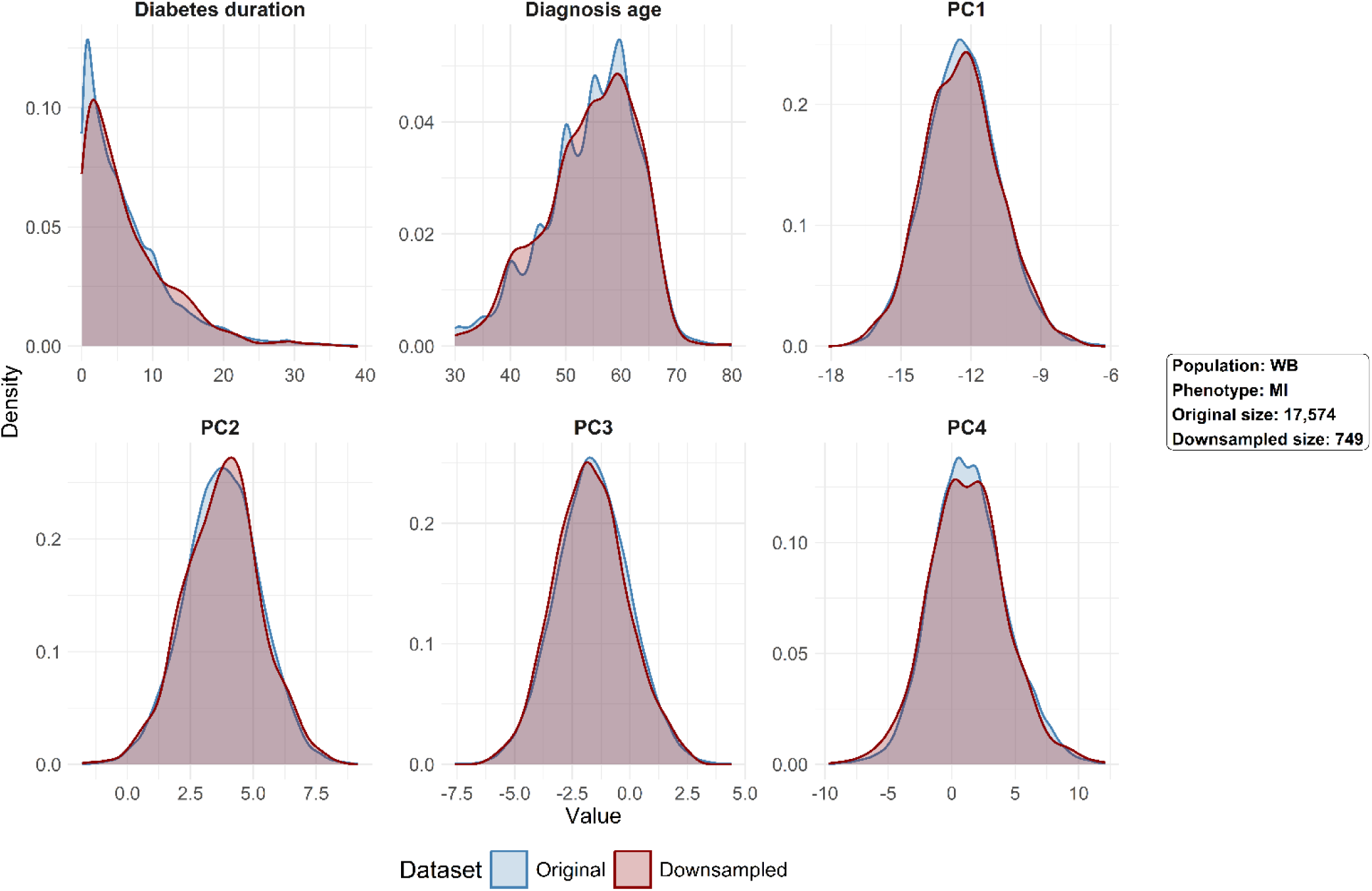
Comparative density plots of continuous covariates used in the predictive models in the White British (**WB**) for myocardial Infarction (**MI**). Distributions are shown for both the original sample and the downsampled dataset. Covariates include age at diabetes diagnosis, diabetes duration, and the first four principal components (**PC1** to **PC4**). Initial distributions were largely preserved after downsampling, indicating good retention of the underlying data structure.

**Supplementary Fig. 6.**
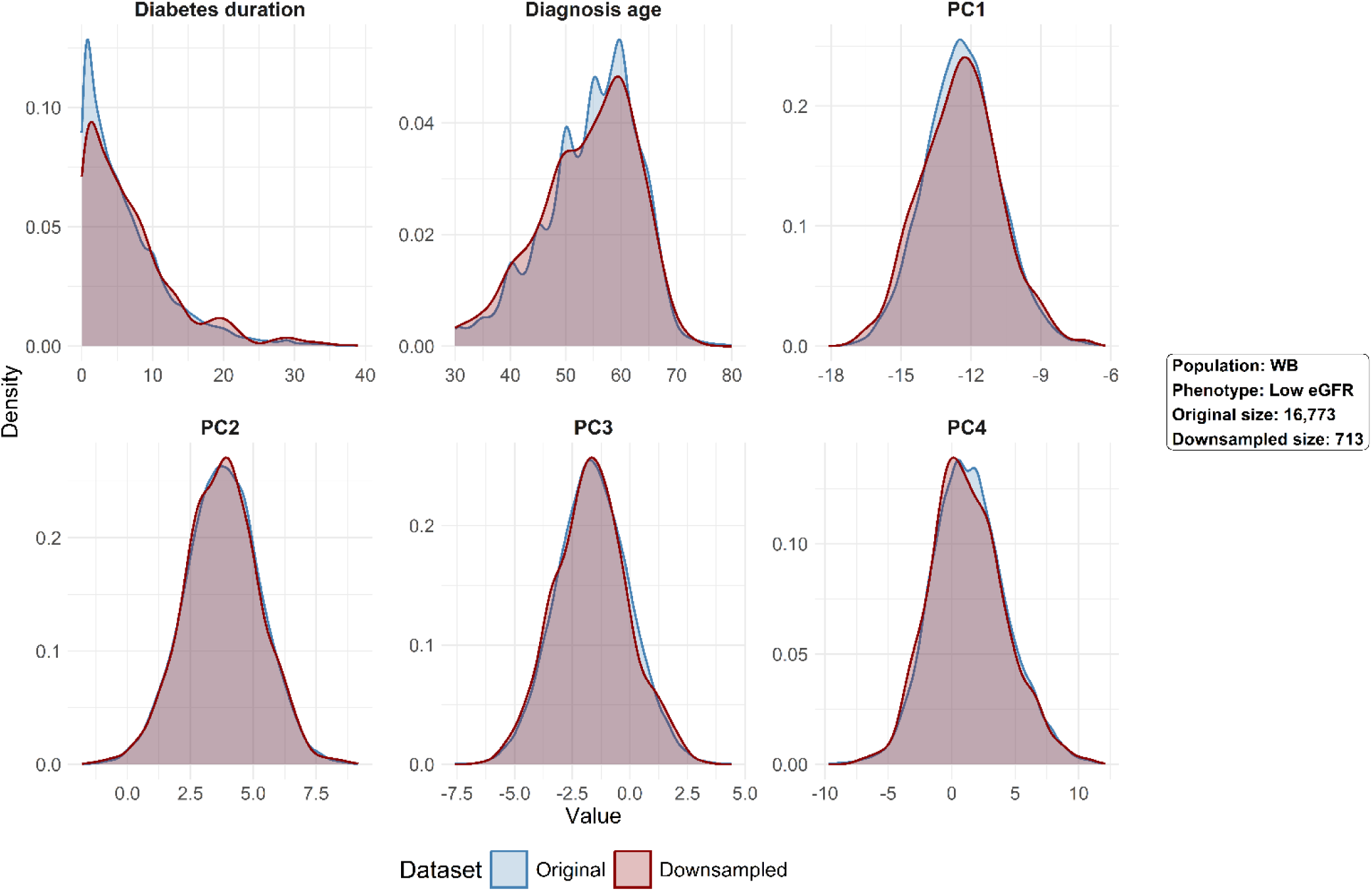
Comparative density plots of continuous covariates used in the predictive models in the White British (**WB**) population for **Low eGFR**. Distributions are shown for both the original sample and the downsampled dataset. Covariates include age at diabetes diagnosis (**age_diag**), diabetes duration (**diab_year**), and the first four principal components (**PC1** to **PC4**). Initial distributions were largely preserved after downsampling, indicating good retention of the underlying data structure.

**Supplementary Fig. 7.**
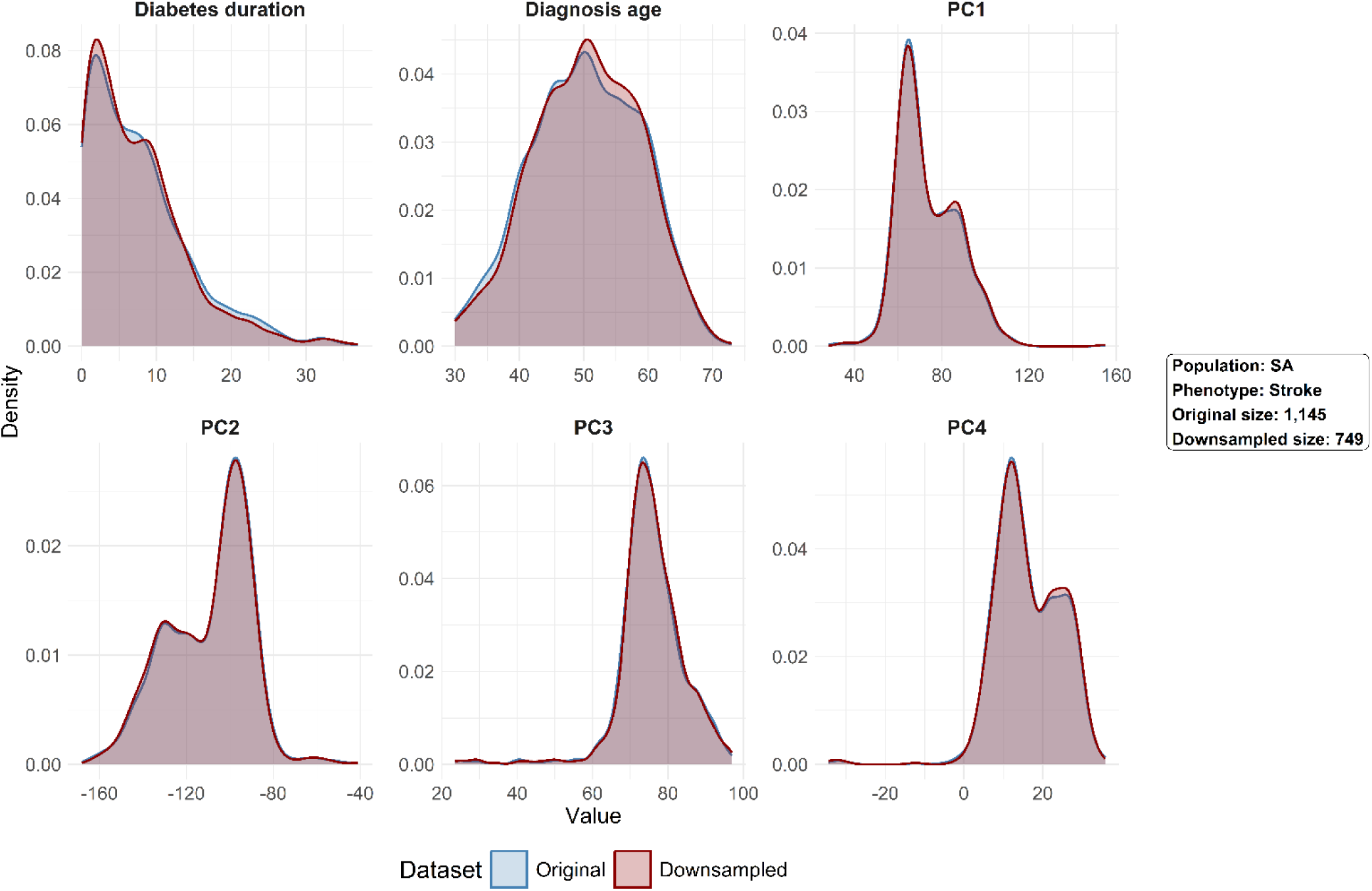
Comparative density plots of continuous covariates used in the predictive models in the South Asian (**SA**) population for **Stroke**. Distributions are shown for both the original sample and the downsampled dataset. Covariates include age at diabetes diagnosis (**age_diag**), diabetes duration (**diab_year**), and the first four principal components (**PC1** to **PC4**). Initial distributions were largely preserved after downsampling, indicating good retention of the underlying data structure.

**Supplementary Fig. 8.**
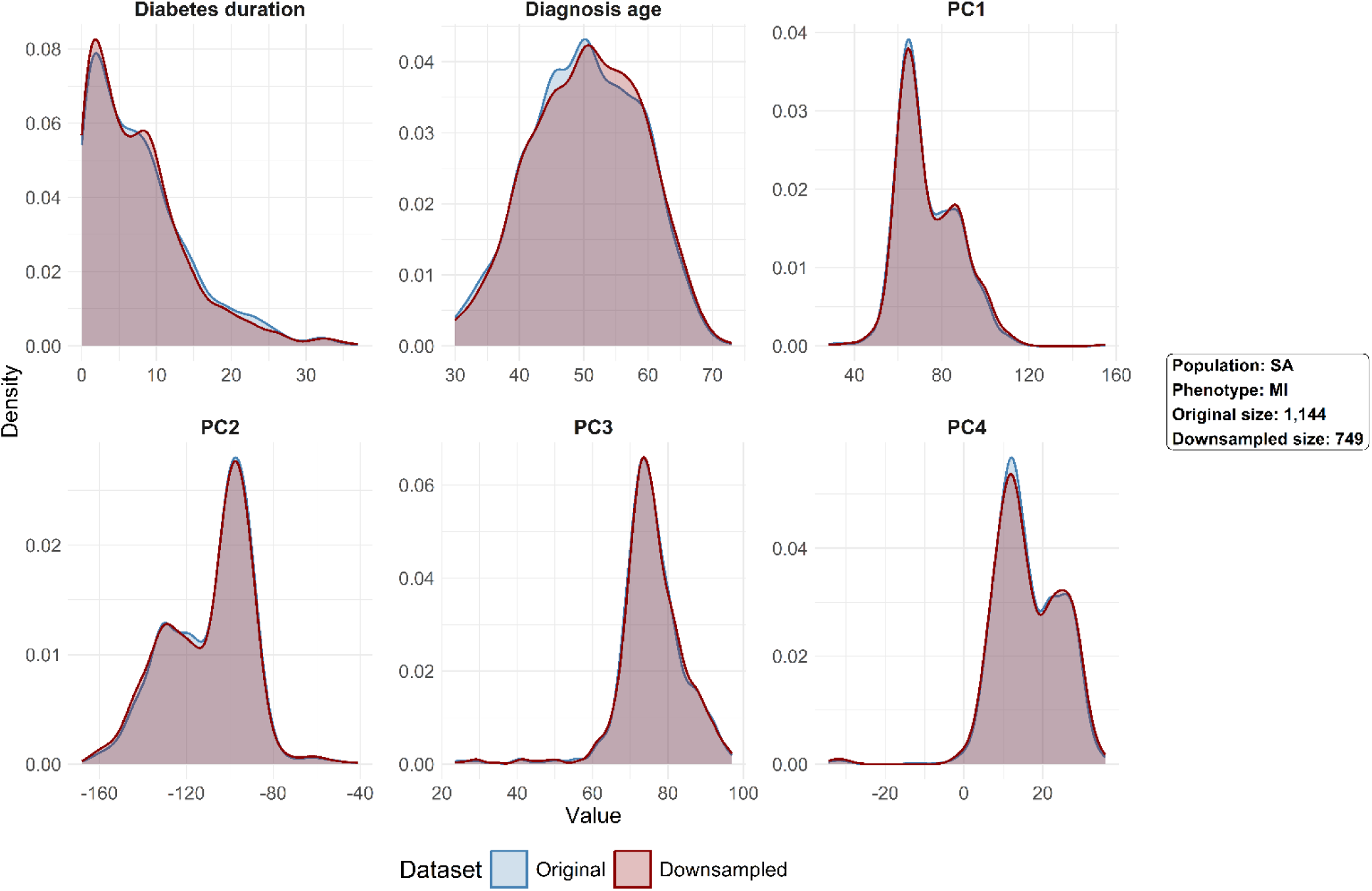
Comparative density plots of continuous covariates used in the predictive models for the South Asian (**SA**) population for myocardial Infarction (**MI**). Distributions are shown for both the original sample and the downsampled dataset. Covariates include age at diabetes diagnosis (**age_diag**), diabetes duration (**diab_year**), and the first four principal components (**PC1** to **PC4**). Initial distributions were largely preserved after downsampling, indicating good retention of the underlying data structure.

**Supplementary Fig. 9.**
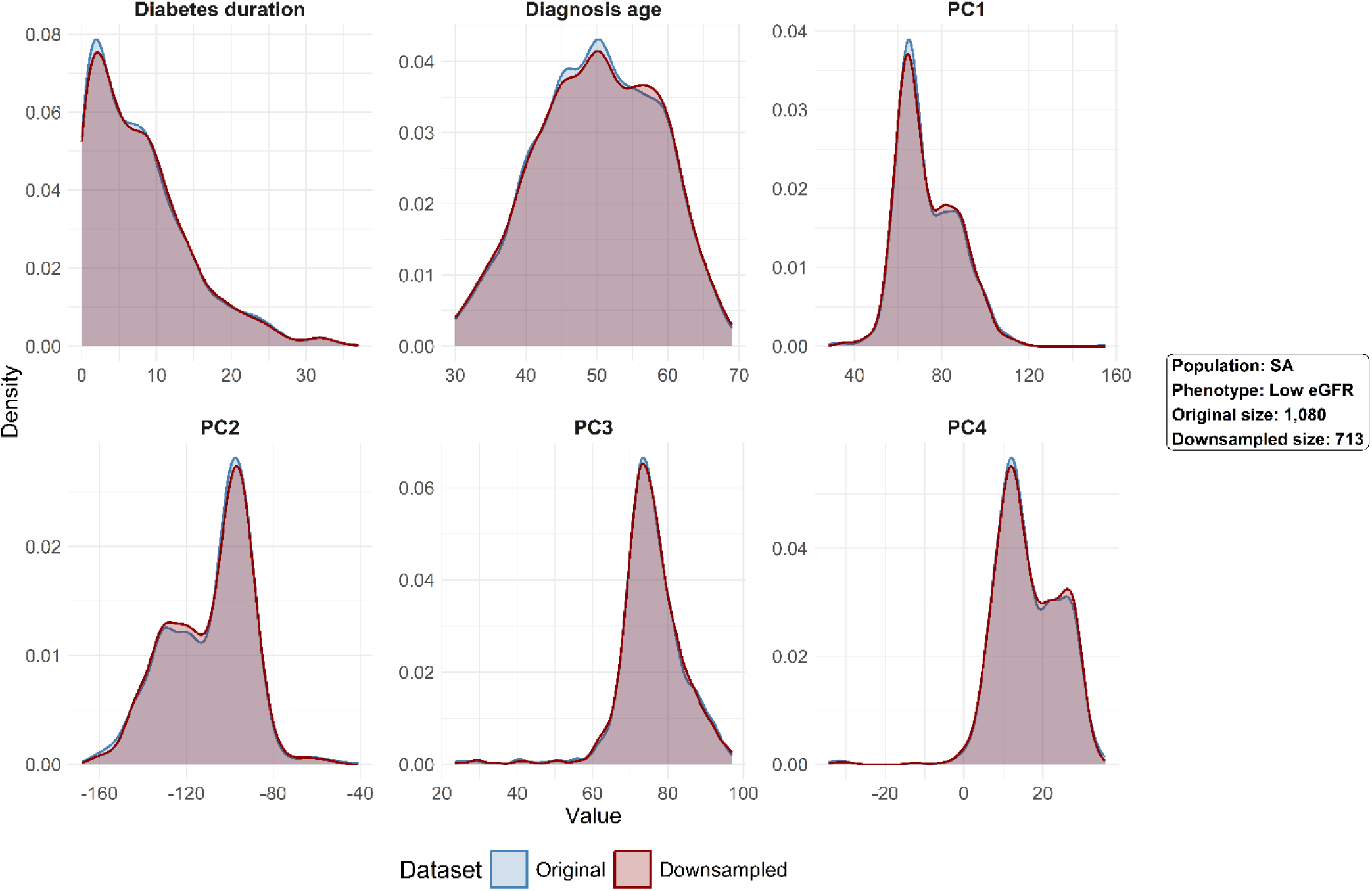
Comparative density plots of continuous covariates used in the predictive models in the South Asian (**SA**) population for **Low eGFR**. Distributions are shown for both the original sample and the downsampled dataset. Covariates include age at diabetes diagnosis, diabetes duration, and the first four principal components (**PC1** to **PC4**). Initial distributions were largely preserved after downsampling, indicating good retention of the underlying data structure.

**Supplementary Fig. 10.**
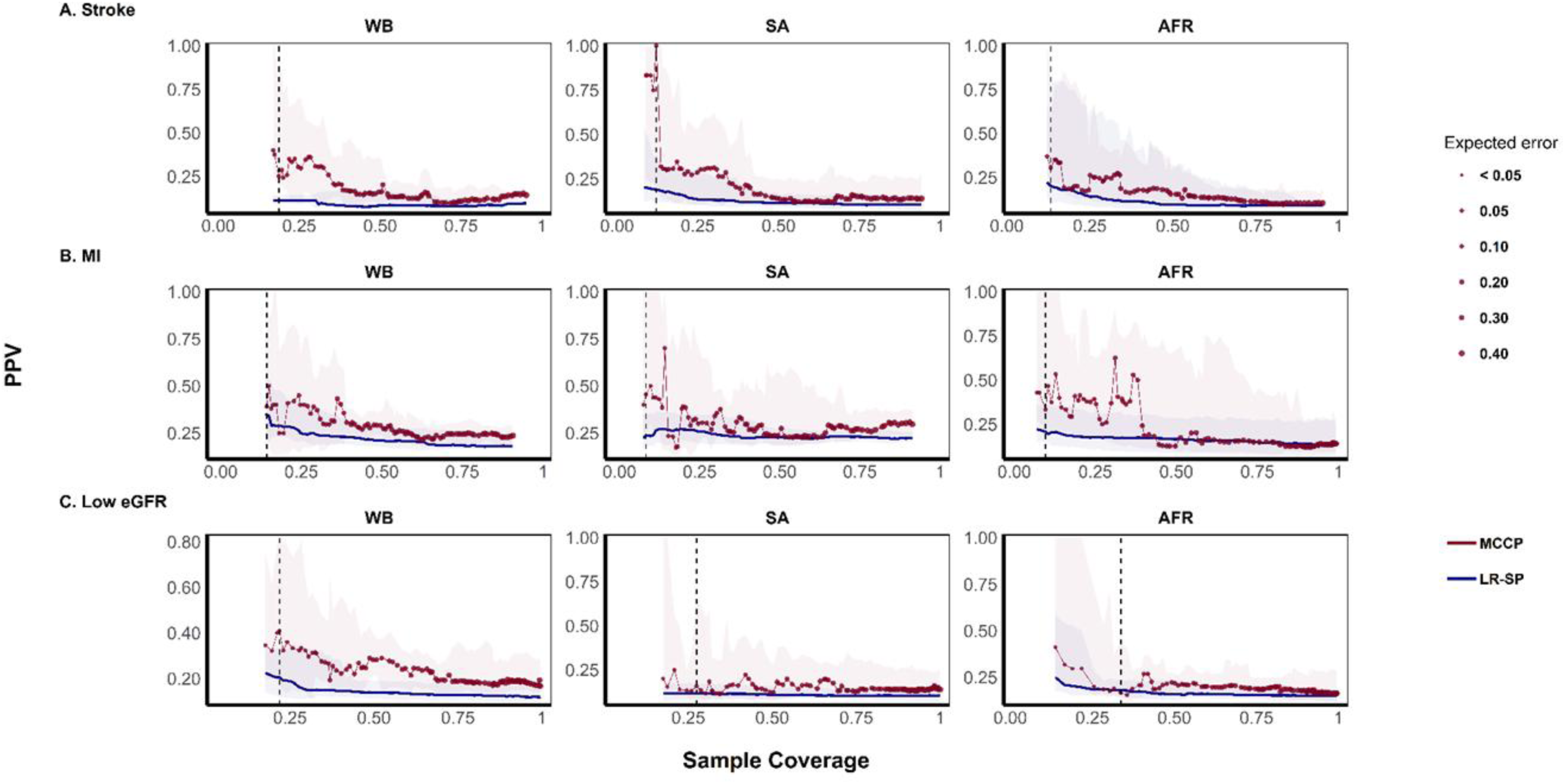
PPV after sample size harmonization. Same as Supplementary Fig. 1, except that the WB and SA populations were downsampled via stratified random subsampling to match the sample size of the AFR population.

**Supplementary Fig. 11.**
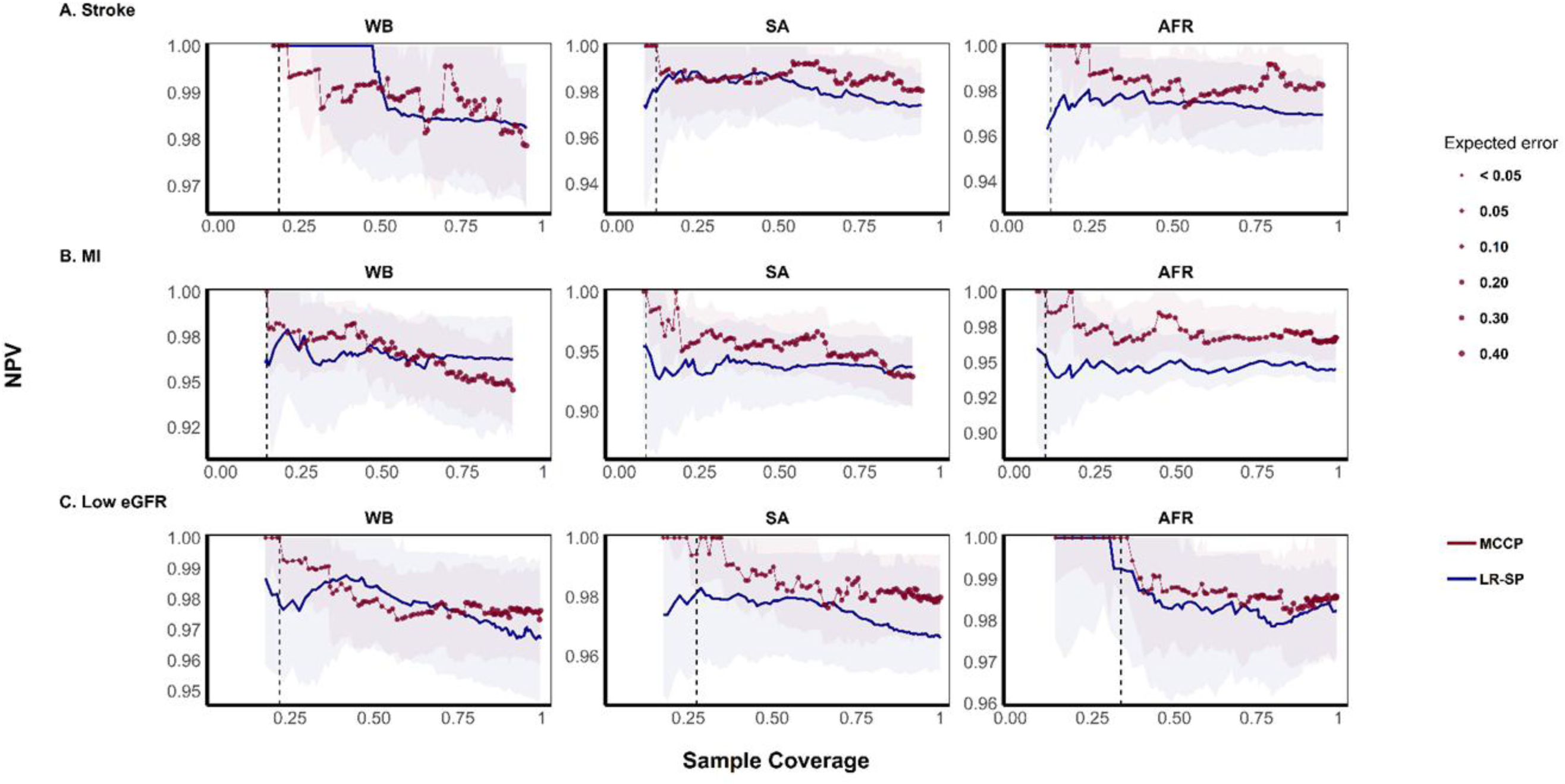
NPV after sample size harmonization. Same as Supplementary Fig. 2, except that the WB and SA populations were downsampled via stratified random subsampling to match the sample size of the AFR population.

**Supplementary Fig. 12.**
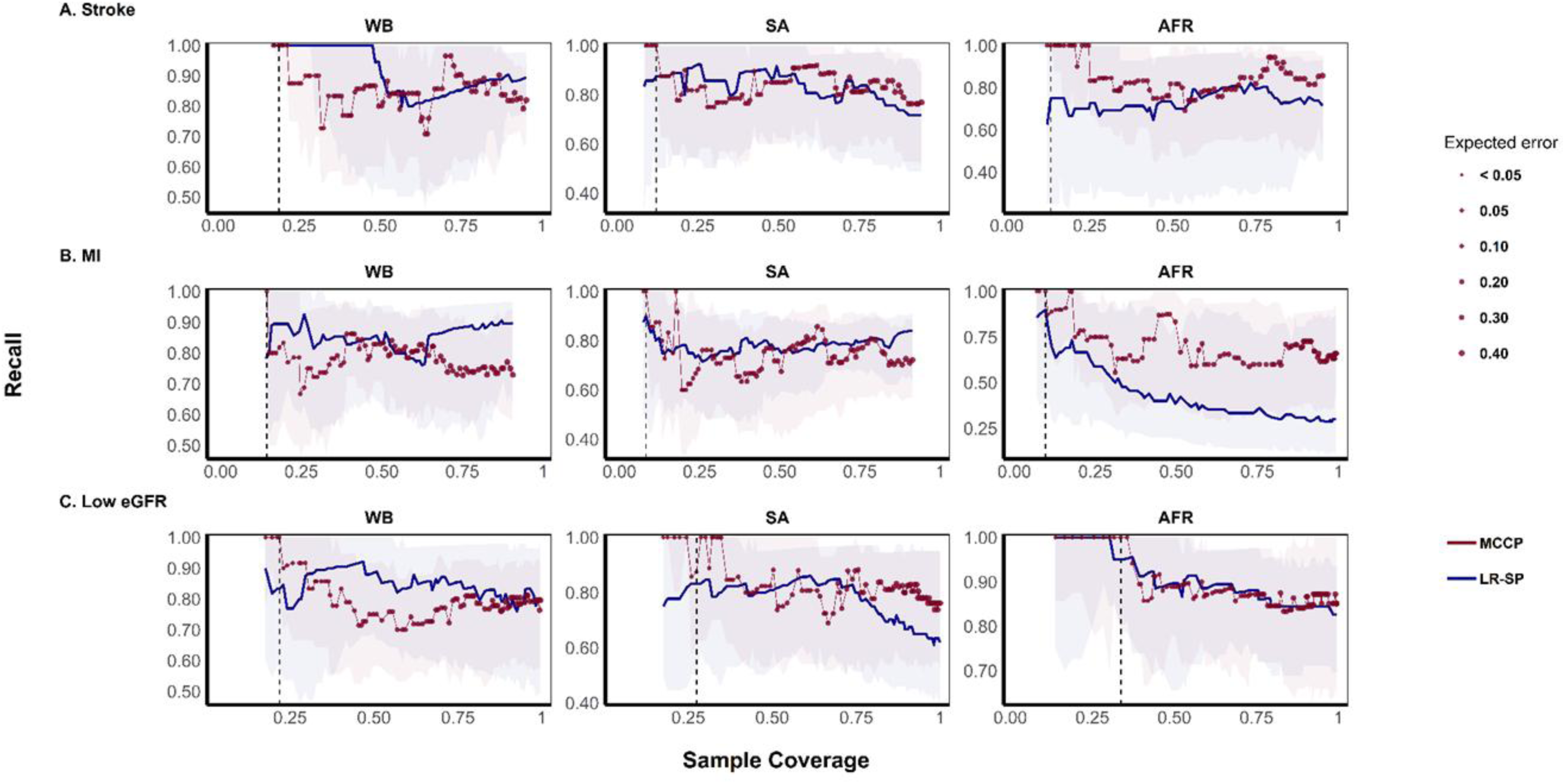
Recall after sample size harmonization. Same as Supplementary Fig. 3, except that the WB and SA populations were downsampled via stratified random subsampling to match the sample size of the AFR population

## References

1. Sun, H., et al., IDF Diabetes Atlas: Global, regional and country-level diabetes prevalence estimates for 2021 and projections for 2045. Diabetes Res Clin Pract, 2022. 183: p. 109119.

2. Chatterjee, S., K. Khunti, and M.J. Davies, Type 2 diabetes. The Lancet, 2017. 389(10085): p. 2239–2251.

3. Zoungas, S., et al., Combined effects of routine blood pressure lowering and intensive glucose control on macrovascular and microvascular outcomes in patients with type 2 diabetes: new results from the ADVANCE trial. Diabetes care, 2009. 32(11): p. 2068–2074.

4. Chalmers, J., et al., Intensive blood glucose control and vascular outcomes in patients with type 2 diabetes. 2008.

5. Tremblay, J., et al., Polygenic risk scores predict diabetes complications and their response to intensive blood pressure and glucose control. Diabetologia, 2021. 64: p. 2012–2025.

6. Martin, A.R., et al., Human Demographic History Impacts Genetic Risk Prediction across Diverse Populations. Am J Hum Genet, 2017. 100(4): p. 635–649.

7. Martin, A.R., et al., Clinical use of current polygenic risk scores may exacerbate health disparities. Nature Genetics, 2019. 51(4): p. 584–591.

8. Need, A.C. and D.B. Goldstein, Next generation disparities in human genomics: concerns and remedies. Trends in Genetics, 2009. 25(11): p. 489–494.

9. Bustamante, C.D., F.M. De La Vega, and E.G. Burchard, Genomics for the world. Nature, 2011. 475(7355): p. 163–165.

10. Petrovski, S. and D.B. Goldstein, Unequal representation of genetic variation across ancestry groups creates healthcare inequality in the application of precision medicine. Genome biology, 2016. 17: p. 1–3.

11. Popejoy, A.B. and S.M. Fullerton, Genomics is failing on diversity. Nature, 2016. 538(7624): p. 161–164.

12. Patel, A., Effects of a fixed combination of perindopril and indapamide on macrovascular and microvascular outcomes in patients with type 2 diabetes mellitus (the ADVANCE trial): a randomised controlled trial. The Lancet, 2007. 370(9590): p. 829–840.

13. Cortés-Ciriano, I. and A. Bender, Concepts and Applications of Conformal Prediction in Computational Drug Discovery, in Artificial Intelligence in Drug Discovery, N. Brown, Editor. 2020, The Royal Society of Chemistry. p. 0.

14. Gammerman, A., et al., Conformal and Probabilistic Prediction with Applications: 5th International Symposium, COPA 2016, Madrid, Spain, April 20-22, 2016, Proceedings. Vol. 9653. 2016: Springer.

15. Sun, J., et al., Applying Mondrian Cross-Conformal Prediction To Estimate Prediction Confidence on Large Imbalanced Bioactivity Data Sets. Journal of Chemical Information and Modeling, 2017. 57(7): p. 1591–1598.

16. Shafer, G. and V. Vovk, A tutorial on conformal prediction. Journal of Machine Learning Research, 2008. 9(3).

17. Sun, J., et al., Translating polygenic risk scores for clinical use by estimating the confidence bounds of risk prediction. Nature Communications, 2021. 12(1): p. 5276.

18. Hamet, P., et al., PROX1 gene CC genotype as a major determinant of early onset of type 2 diabetes in slavic study participants from Action in Diabetes and Vascular Disease: Preterax and Diamicron MR Controlled Evaluation study. J Hypertens, 2017. 35 **Suppl 1**(Suppl 1): p. S24–s32.

19. Sudlow, C., et al., UK biobank: an open access resource for identifying the causes of a wide range of complex diseases of middle and old age. PLoS medicine, 2015. 12(3): p. e1001779.

20. Ali, M., PyCaret: An open-source, low-code machine learning library in Python. 2020.

21. Vovk, V., A. Gammerman, and G. Shafer, Algorithmic learning in a random world. Vol. 29. 2005: Springer.

22. Levey, A.S., et al., A new equation to estimate glomerular filtration rate. Ann Intern Med, 2009. 150(9): p. 604–12.

23. Conover, W.J., Practical nonparametric statistics. 1999: john wiley & sons.

24. Agresti, A., Categorical data analysis. 2013: John Wiley & Sons.

25. Duncan, L., et al., Analysis of polygenic risk score usage and performance in diverse human populations. Nature Communications, 2019. 10(1): p. 3328.

26. Gomez, F., J. Hirbo, and S.A. Tishkoff, Genetic variation and adaptation in Africa: implications for human evolution and disease. Cold Spring Harbor perspectives in biology, 2014. 6(7): p. a008524.

27. Kamiza, A.B., et al., Transferability of genetic risk scores in African populations. Nature Medicine, 2022. 28(6): p. 1163–1166.

28. Scutari, M., I. Mackay, and D. Balding, Using genetic distance to infer the accuracy of genomic prediction. PLoS genetics, 2016. 12(9): p. e1006288.

29. Martin, A.R., et al., Human Demographic History Impacts Genetic Risk Prediction across Diverse Populations. The American Journal of Human Genetics, 2017. 100(4): p. 635–649.

30. Kachuri, L., et al., Principles and methods for transferring polygenic risk scores across global populations. Nature Reviews Genetics, 2024. 25(1): p. 8–25.

31. Zhou, C., et al., The nonlinear relationship between estimated glomerular filtration rate and cardiovascular disease in US adults: a cross-sectional study from NHANES 2007-2018. Front Cardiovasc Med, 2024. 11: p. 1417926.

32. Ahmed, S., et al., Examining the Potential Impact of Race Multiplier Utilization in Estimated Glomerular Filtration Rate Calculation on African-American Care Outcomes. Journal of General Internal Medicine, 2021. 36(2): p. 464–471.

33. Eneanya, N.D., W. Yang, and P.P. Reese, Reconsidering the Consequences of Using Race to Estimate Kidney Function. Jama, 2019. 322(2): p. 113–114.

34. Udler, M.S., et al., Effect of Genetic African Ancestry on eGFR and Kidney Disease. J Am Soc Nephrol, 2015. 26(7): p. 1682–92.

35. Inker, L.A., et al., New Creatinine– and Cystatin C–Based Equations to Estimate GFR without Race. New England Journal of Medicine, 2021. 385(19): p. 1737–1749.

36. Horimoto, A., D. Xue, and J. Cai, Genome-Wide Admixture Mapping of Estimated Glomerular Filtration Rate and Chronic Kidney Disease Identifies European and African Ancestry-of-Origin Loci in Hispanic and Latino Individuals in the United States. 2022. 33(1): p. 77–87.

37. Parikh, R., et al., Understanding and using sensitivity, specificity and predictive values. Indian J Ophthalmol, 2008. 56(1): p. 45–50.

38. Trevethan, R., Sensitivity, specificity, and predictive values: foundations, pliabilities, and pitfalls in research and practice. Frontiers in public health, 2017. 5: p. 307.

39. Fatumo, S., et al., A roadmap to increase diversity in genomic studies. Nature Medicine, 2022. 28(2): p. 243–250.

40. Corpas, M., et al., Bridging genomics&#x2019; greatest challenge: The diversity gap. Cell Genomics, 2025. 5(1).

